# Biological Therapy for Chronic Obstructive Pulmonary Disease (COPD): Efficacy and Safety

**DOI:** 10.1101/2025.10.17.25337825

**Authors:** Olga Milena García Morales, Ariadna Auladell, Daniel Poloni, Luz Angela Torres-López, Laura Díaz, Gerard Urrútia Couchi, María Ximena Rojas

## Abstract

**Background:** The predominant airway inflammation in chronic obstructive pulmonary disease (COPD) is neutrophilic, although numerous studies have shown that eosinophilic inflammation contributes significantly to COPD. Eosinophilic COPD patients are characterized by a greater number of total exacerbations and higher hospitalization rates.

Biological therapy targets multiple steps in eosinophilic inflammation, providing a promising approach to reduce exacerbation frequency and mortality.

**Objective:** To provide a timely, rigorous, and continuously updated summary of the evidence available on the role of biological therapy for the treatment of patients with COPD.

**Design:** This is a Living Evidence synthesis, that starts with a baseline synthesis report of the effects of the intervention on the main predefined outcomes. Based on its conclusions, we will set up the living evidence approach, and the evidence monitoring will begin.

**Methods:** *Evidence identification, screening, and selection:* Automated living searches were performed in relevant databases following the Epistemonikos procedures. Results were incorporated into the Epistemonikos “Living OVerview of Evidence (L.OVE)” platform which was used for evidence screening, and selection. Two reviewers screened all titles and abstracts supported by the platform automated classifiers that excluded references with a low probability of being relevant. For this baseline report we included randomized trials that evaluated the use of biological therapy compared to the use of standard-of-therapy alone in adult patients with COPD. Main outcomes defined as critical or important for decision making include exacerbation rate, exacerbation-free time, lung function and quality of life. Two reviewers independently screened each study for eligibility, extract data, and assess its methodological quality using appropriate tools. We performed meta-analyses of the study’s results when pertinent. We applied the GRADE approach to assess the certainty of the evidence found for each outcome. We will continuously monitor the evidence by performing daily searches and monthly screening of the retrieved references. Additionally, each three months we will manually search for ongoing studies in the International Clinical Trials Registry Platform trial registries. The evidence monitoring, including decisions to incorporate evidence and withdraw the question from the living mode will follow the process proposed by the Living Evidence to Inform Health Decisions framework (ref) as stated in our protocol. A living, web-based version of this review will be openly available during the next year at https://livingevidenceihd.com/lesrepo/. We will resubmit it every time the conclusions change or whenever there are substantial updates.

**Results:** Initial searches retrieved 24 systematic reviews, 2 evaluating the use of biologics on COPD, which included 6 randomized control trials. Additional searches retrieved 125 randomized controlled trials (RCTs). Of these, 31 RCTs were deemed potentially eligible and were reviewed in full text. Ultimately, 9 RCTs evaluating the use of biological therapies (MEDI8968, benralizumab, mepolizumab, dupilumab, astegolimab, and itepekimab) compared to placebo in COPD patients were included. The GRADE quality of evidence for the main outcomes was assessed as intermediate.

**Conclusions:** The evidence suggests that biological therapies may reduce the risk of exacerbations in patients with chronic obstructive pulmonary disease (COPD). However, the overall impact on lung function and quality of life remains inconclusive. The variability in study outcomes, particularly concerning SGRQ scores and FEV1, emphasizes the need for continuing monitoring evidence.

**PROSPERO /OSF**

**Protocol Registration DOI:** 10.17605/OSF.IO/SUYD2

**STRUCTURED ABSTRACT:** *Objective:* To synthesize and update evidence on the efficacy and safety of biological therapy for chronic obstructive pulmonary disease (COPD).

*Methods:* This living evidence synthesis included systematic reviews (SRs) and randomized controlled trials (RCTs) comparing biological therapies (e.g., benralizumab, mepolizumab, dupilumab) with placebo or standard care in adults with moderate to severe COPD. Automated searches were conducted using the Epistemonikos “Living Overview of Evidence” (L.OVE) platform, supplemented by manual searches. Two reviewers independently screened studies, extracted data, and assessed risk of bias using Cochrane RoB2 for RCTs and AMSTAR II for SRs. Meta-analyses were performed, and the certainty of evidence was evaluated using the GRADE approach.

*Results:* Nine RCTs were included. Biological therapy probably reduces annual exacerbation rates (moderate certainty; MD -0.12, 95% CI -0.23 to 0.00). The evidence is very uncertain about its effect on quality of life (SGRQ score: MD 0.05, 95% CI -0.04 to 0.14; very low certainty) due to serious inconsistency (I² = 95.5%) and imprecision. Biological therapy may result in little to no difference in lung function (FEV1: MD 0.02, 95% CI -0.02 to 0.06; low certainty), with serious inconsistency (I² = 76.8%) and imprecision. Biological therapy probably results in no difference in total adverse events (moderate certainty; RR 1.02, 95% CI 0.99 to 1.05) but likely reduces severe adverse events (high certainty; RR 0.84, 95% CI 0.77 to 0.93).

*Conclusions:* Current evidence suggests that biological therapies for COPD provide little to no clinical benefit over standard care in terms of exacerbation rates, lung function, or safety, with uncertain effects on quality of life. Biological therapy probably has little to no effect on annual exacerbations compared with standard care. There is likely no meaningful difference in lung function (FEV1) compared with standard care. Biologicals probably result in no difference in the number of total adverse events. Regarding the effect on quality of life, measured by the SGRQ compared with standard care, the evidence is very uncertain; therefore, no conclusions can be drawn. Overall, the certainty of the evidence supporting these statements is generally moderate, but is particularly limited by imprecision in estimates when considering thresholds for the minimal important difference (MID).

## Introduction

Chronic obstructive respiratory disease (COPD) is a leading cause of mortality, morbidity and economic burden. In 2019, COPD accounted for 3,3 million deaths and 213.3 million prevalent cases reported globally. (1,2)Chen et al estimated that in 2020 the overall cost of COPD was INT$4·326 trillion, approximately 51.796 billion dollars, highlighting its widespread impact (3)

Multiple factors contribute to mortality in COPD, including age and cardiovascular comorbidities. However, among these factors, exacerbations emerge as a paramount predictor of mortality. Studies have demonstrated that patients experiencing two or more exacerbations face a poor prognosis, with a mortality rate of 31% at 6 months (4). In contrast, those without exacerbations exhibit a significantly higher survival rate of 80% over the same period. (5) Furthermore, compared to the general population, patients with COPD following a severe exacerbation have a lower mean life expectancy. Van Hirtum et al. estimated that COPD patients had a mean life expectancy of 9.7 years versus 10.2 years for the general population.(6) A significant negative prognostic factor for these patients is the frequency of exacerbations, which adversely impacts overall survival.(6)

The COPD is characterized by persistent airflow limitation and respiratory symptoms due to airway/alveolar abnormalities and associated chronic inflammation that worsens the structural damages. The pattern of inflammation present in COPD can vary, with the most common one being predominated by neutrophils, cytotoxic CD8+ T cells, and alveolar macrophages. However, in some patients, eosinophils play a significant role in inflammation. (7,8) Once activated by IL5 and having migrated to the lungs, eosinophils release multiple proinflammatory chemokines, including IL13, which promotes alveolar macrophages and exacerbates alveolar damage.

Biological therapy is a promising treatment approach for COPD that aims to reduce inflammation, thereby reducing the risk of exacerbation and improving patientś quality of life and prognosis. There are new monoclonal antibodies that target eosinophilic activation and proliferation. This target includes IL-33, IL-5 (main chemokine for eosinophil activation), anti-IL-5 receptor antagonist, anti IL4/13 receptor and ST2 (IL1 receptor-like 1).

Given the recent use of these types of therapies in the treatment of COPD, it is expected that new evidence will emerge regarding their effects on these patients, as well as research on the development of new technologies. This approach will make it possible to develop a rigorous and updated synthesis of the evidence on the risk of exacerbation, lung function and quality of life and draw conclusions on the impact of biological therapy in those aspects.

This living evidence synthesis is being developed as part of the Living Evidence to Inform Health Decisions Program (8), which supports health system organizations, guideline development groups and other institutions in the implementation of a living process for the development of the synthesis of the evidence to inform health decisions. The continues update resulting from evidence surveillance will be available at program website (https://livingevidenceframework.com/)

## Methods

### Protocol and registration

This is a baseline synthesis report of a Living Evidence Synthesis (LES) of the effects of biological therapy on the COPD exacerbation rate. Its design and planning comply with the Methods for Planning and Reporting Living Evidence Synthesis checklist proposed by Bendersky et al.(10) A generic protocol stating the methodology of living evidence syntheses to be conducted within the LE-IHD program projects has been published elsewhere, and adapted or this evidence synthesis / systematic review to the specificities of the question and registered in OFR osf.io/edtfj

This manuscript complies with the ‘Preferred Reporting Items for Systematic reviews and Meta-Analyses’ (PRISMA) guidelines (6) (See Appendix 1).

#### Evidence identification, screening and selection

Living searches were performed from inception in all databases following the Epistemonikos procedures. No date, publication status, or language restriction were applied to the searches. The Epistemonikos “Living OVerview of Evidence” (L.OVE) platform (11) was used as a technological enabler for evidence screening, and selection. Authors worked with the Epistemonikos team maintaining the L·OVE platform in devising the literature search aimed to identify initially systematic reviews and subsequent randomized clinical trials.

For this baseline report, the evidence identified and retrieved to the L·OVE of this question (<u>Chronic obstructive pulmonary</u> disease until [30/July/2023] was screened by two researchers independently. We assessed the title and abstract against the inclusion criteria and obtained the full reports for all titles that appeared to meet the inclusion criteria or required further analysis to decide about their inclusion.

We included randomized clinical trials assessing biological therapy vs placebo in adult patients with moderate to severe COPD. We did not restrict our selection criteria to any dosage or route of administration. We did not use the outcomes as an inclusion criterion during the selection process.

#### Extraction and management of data

Using standardized forms, one reviewer extracted data from each study included. We collected information related to the study design, characteristics of participants (including disease severity and age), study eligibility criteria and details of the intervention. The outcomes assessed (COPD exacerbation rate, change in SGRQ score and FEV1 changes) and the time they were measured, the source of funding of the study and the conflicts of interest disclosed by the investigators. For systematic reviews we collected the risk of bias assessment for each individual study reporting on the outcomes of interest for this review as well as the pull estimate of the effect reported for each important outcome. For RCTs we collected the data necessary to estimate the effect measure of our interest according to the type of variable e.g. dichotomous or continues) (see measurement of treatment effect).

#### Risk of bias assessment

We assessed the risk of bias of the whole evidence we identified in searches using appropriate instruments according to the type of study: AMSTAR II tool for SRs [30], and Cochrane RoB 2 tool for RCTs [31]. In the case of including NRS, these will be evaluated according to the ROBINS-I tool [32]. Two reviewers will independently assess all the included studies.

Discrepancies between review authors were resolved by discussion to reach consensus. If necessary, a third review author was consulted to achieve a decision.

#### Measures of treatment effect

The main outcomes of interest were the annualized rate of COPD exacerbations, change from baseline in prebronchodilator FEV1 and change from baseline in St. George’s Respiratory Questionnaire (SGRQ) score.

For dichotomous outcomes, we expressed the estimate of treatment effect as (hazard ratio (HR), risk ratios (RR) or odds ratios (OR) along with 95% confidence intervals (CI).

For continuous outcomes, we used mean difference and standard deviation (SD) to summaries the data using a 95% CI.

We considered the following factors as baseline potential confounders: smoking status.

#### Data synthesis

We performed meta-analyses using RevMan 5 with a using the inverse variance method with random effects model.

#### Subgroup and sensitivity analysis

We planned to perform subgroup analysis according to eosinophilic count, we consider three subgroups of patients based on blood eosinophil count, following the 2023 GOLD guidelines: <100 cells/uL, 100-300 cells/uL, and >300 cells/uL. Based on the baseline blood eosinophil count, we have included studies within each of these ranges for three outcomes: exacerbation rate, patients with exacerbations, and mortality. In case we identified significant differences between subgroups (test for interaction <0.05) we considered to report the results of individual subgroups separately.

#### Assessment of certainty of evidence

We judged the certainty of the evidence for all outcomes using the Grading of Recommendations Assessment, Development and Evaluation working group methodology (GRADE Working Group)(12) across the domains of risk of bias, consistency, directness, precision, and reporting bias. Certainty was adjudicated as high, moderate, low or very low. For the main comparisons and outcomes, we prepared a Summary of Findings (SoF) table (13,14)

#### Living evidence synthesis

To maintain the living evidence process for this review, we will keep a living search to detect randomized controlled trials and NRS. An artificial intelligence algorithm deployed in the L·OVE <u>Chronic obstructive pulmonary</u> will provide instant notification of articles with a high likelihood to be eligible. Additionally, each three months, we will manually search for ongoing studies in the WHO International Clinical Trials Registry Platform and the clinicaltrials.gov.

One reviewer will be in charge of assessing the evidence that has entered the L.OVE of this question every month and apply the selection criteria presented above. If a potentially eligible study is found, a second reviewer will confirm its eligibility by reading the full text. Results of evidence surveillance will be collected and keep as part of the study records. Information on PRISMA will be updated accordingly. Criteria for selecting studies will be revised and changed accordingly during the LE processes each 4 months.

All new eligible studies will undergo a data extraction process. The data synthesis will be updated immediately after that considering the predefined subgroups of interest, and the body of evidence for the outcomes of interest will be assessed following the GRADE approach accordingly looking for changes on the certainty assessment results.

#### Statistical considerations for the living evidence synthesis

As part of evidence surveillance, the inclusion of new studies identified reporting on the outcomes of interest will follow this approach: We will perform a meta-analysis for each of the outcomes of interest reported by the new studies using a fixed-effect model to evaluate the statistical heterogeneity among included studies by using the I statistics. If new heterogeneity is detected (i.e. increase the heterogeneity previously identified or new heterogeneity arises where it was previously undetected), we will explore its potential sources by reviewing the new studies against previously included studies in order to identify reasons that may explain inconsistent results among studies. In presence of unexplained heterogeneity (I2> 70%), we will consider not to Meta-analyze them and explain the evidence synthesis narratively. If the I is below 90%, we will perform a meta-analysis by using the fixed effects of the random effects model, whichever pertinent.

## Results

### Results of the search

The figure 1 summarizes the searching results. The search retrieved 8 SRs and 125 RCTs to the L·OVE platform. Two SRs evaluated the use of biological therapy in chronic obstructive pulmonary disease (COPD). However, due to their limited scope, restricted exclusively to IL-5 therapies and the timing of their publication—while important randomized controlled trials were ongoing—these reviews were excluded from the baseline synthesis.

**Figure 1.**
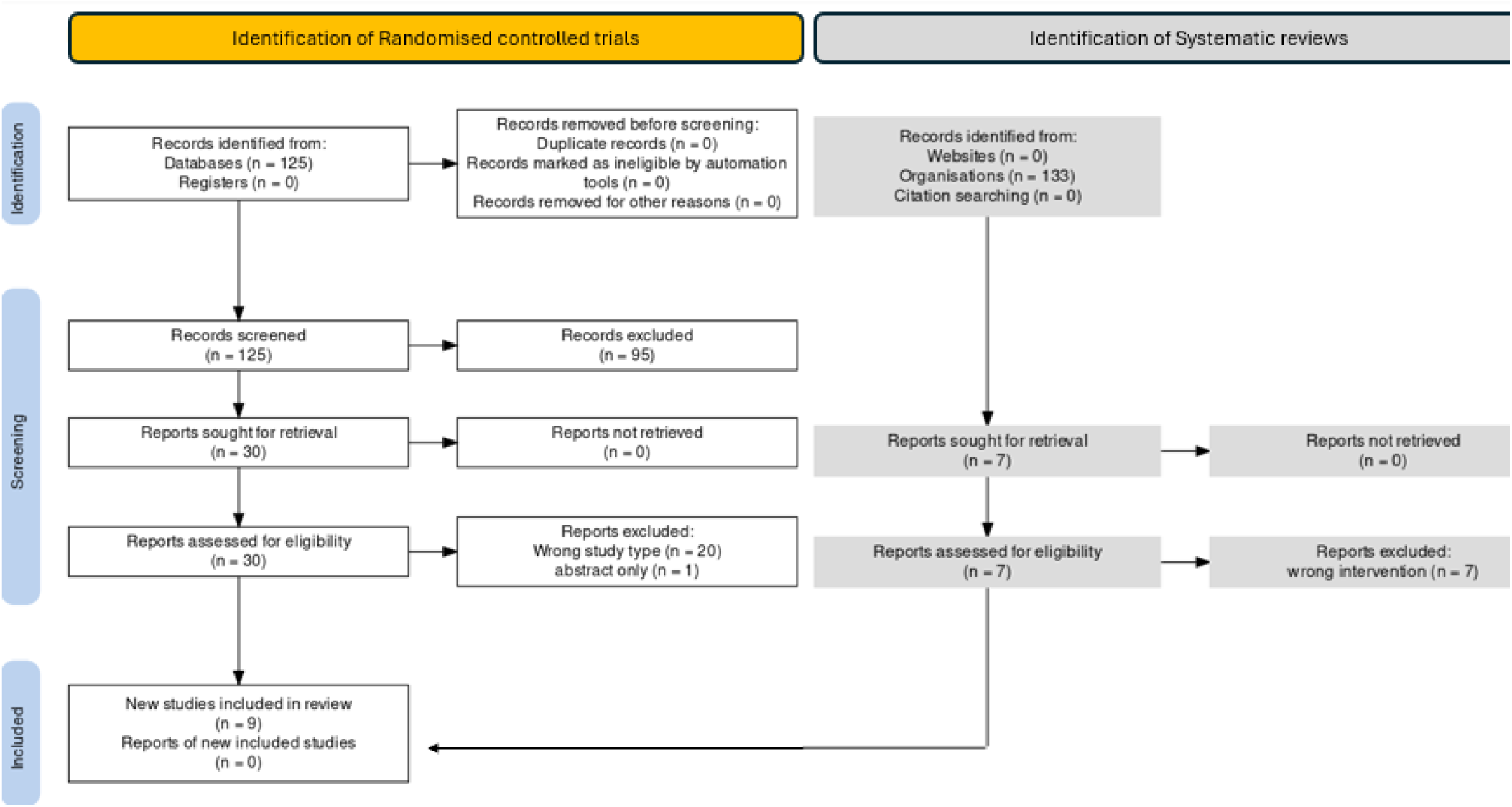
PRISMA Flowchart.

We considered 31 RCTs as potentially eligible that were retrieved and evaluated in full texts. A total of 9 RCT met the inclusion criteria from which we performed the data extraction and conducted meta analysis for the main outcomes (COPD exacerbation rates, change in baseline in SGRQ score and change in baseline FEV1 percent of predicted).

### Characteristics of the included studies

**Yousuf (2022)** is a randomized, doble-blind, placebo–controlled study, comparing the safety and effectiveness of astegolimab at a dosage of 490mg administered every 4 weeks for 44 weeks vs usual care masked with placebo. This study included 81 participants (42 in a comparison of interest in this review) older than 40 years of age, with clinical diagnosis of moderate to severe COPD. Measures of effectiveness included exacerbation rate, change in quality of life (from baseline in St. George’s Respiratory Questionnaire (SGRQ) score), adverse events and change from baseline in post the FEV1 bronchodilator

**Bhatt** (**2023)** is a randomized phase 3, double-blind, placebo-controlled study. The aim was to evaluate the effectiveness and safety of Dupilumab at a dosage of 300mg administered every 2 weeks for 52 weeks compared to standard care masked with placebo. This RCT comprised 939 patients aged 40 to 80 years with a clinical diagnosis of moderate to severe COPD and current or former tobacco smokers who had a smoking history of at least 10 pack-years, with 468 randomly assigned to the intervention group. The measures included exacerbation rate, change in quality of life (from baseline in the SGRQ score), and adverse events.

**METREO (2017),** was a doble-blind, phase 3, placebo-controlled study aimed to compare the effectiveness and safety of two different doses of Mepolizumab 100mg or 300mg administered every 4 weeks for 52 weeks vs usual care masked with placebo. A total of 675 adults, older than 40 years old with moderate-to-severe COPD, who were current smokers, nonsmokers or former smokers, were randomly assigned to any of the three arms. The measures included exacerbation rate, change in quality of life (from baseline in the SGRQ score), and adverse events.

**METREX (2017**), a phase 3, placebo-controlled, doble-blind study evaluated the effectiveness and safety of mepolizumab, a dosage of 100mg administered every 4 weeks subcutaneously, compared to usual care masked with placebo. A total of 837 patients were randomized into four groups based on eosinophilic count: placebo non-eosinophilic (230 patients), mepolizumab with eosinophilic phenotype (233 patients), mepolizumab non-eosinophilic (184 patients), and placebo eosinophilic (190 patients). Participantes were adults with a diagnosis of COPD and ≥2 moderate exacerbations or one severe exacerbation. The measured outcomes included exacerbation rate, change from baseline SGRQ score and adverse events.

**Calverly (2017)** conducted a phase 2, double-blind, placebo-controlled study to compare MEDI8968, administered at a dosage of 300mg every 4 weeks after a 600mg IV loading dose, in terms of safety and efficacy versus standard care plus placebo for 52 weeks. This RCT included patients with a diagnosis of Chronic Obstructive Pulmonary Disease aged 45-75 years and a history of ≥2 exacerbations. Efficacy outcomes were exacerbation rate, adverse events, and change from baseline in postbronchodilator FEV1.

**Rabe (2021)** was double-blind, phase 2a trial aiming to compare Itepekimab, administered as two subcutaneous injections of 150mg every 2 weeks for 52 weeks versus placebo plus standard therapy in terms of efficacy and safety. It included 343 patients aged 40-75 years, former or current smokers, with a diagnosis of COPD at high risk of exacerbations, with 172 randomized to the Itepekimab group. The measured outcomes of interest were exacerbation rate, adverse events, and change from baseline in postbronchodilator FEV1.

**The GALATHEA trial (2019),** was a phase 3, doble-blind, placebo-controlled study, that compared Benralizumab, administered subcutaneously every 4 weeks at two distinct dosages: 30mg or 10mg for 52 weeks to placebo plus standard care in effectiveness and safety profile. It enrolled participants aged 40 to 85 years diagnosed with moderate or severe COPD. Outcomes of interest included the exacerbation rate, change from baseline in the St. George’s Respiratory Questionnaire (SGRQ), and changes in postbronchodilator FEV1 from baseline.

**The TERRANOVA trial (2019),** a doble blind, phase 3, randomized, placebo-controlled study, that evaluated the effectiveness and safety of Benralizumab compared to placebo plus usual care. Benralizumab was administered in different dosage: 100mg, 30mg or 10mg subcutaneously every 4 weeks for 52 weeks. It enrolled 1545 individuals, aged 40 to 85, diagnosed with COPD and a history of frequent moderate or severe exacerbations. Key outcomes of effectiveness included the exacerbation rate, change in the St. George’s Respiratory Questionnaire (SGRQ) score from baseline, changes in postbronchodilator FEV1 from baseline and adverse events.

**Brightling (2014)** conducted a randomized, placebo-controlled, double-blind, phase 2a trial evaluating Benralizumab efficacy and safety compared to usual care plus placebo. Benralizumab was administered as 3 doses of 100mg subcutaneously every 4 weeks, followed by five doses of 100mg every 8 weeks in 101 adult patients aged 40 to 85 years with a diagnosis of moderate-to-severe Chronic Obstructive Pulmonary Disease. The measured outcomes of efficacy were exacerbation rate, change from baseline SGRQ score, and change from baseline in postbronchodilator FEV1. Safety outcomes included adverse events.

Further details and characteristics of the studies can be found in the Annex 2.

### Ongoing studies

We identified 7 ongoing studies, all randomized trials. See Appendix 3 - List of included, excluded and ongoing studies.

### Excluded studies

We excluded 23 RCT that did not fulfill our eligibility criteria. A detailed list of excluded studies with reasons for exclusion is presented in Appendix 2 - List of included, excluded and ongoing studies.

### Risk of bias

Utilizing the ROB-2 assessment tool, all studies demonstrated low risk in the selection of reported results and in the measurement of outcomes. However, Yousuf (2022) had an unclear risk of missing outcome data due to dropouts. Selection bias was low in all studies, except for Brightling (2014) that exhibited unclear bias due to incorrect criteria in patient selection. No other potential sources of bias were identified.

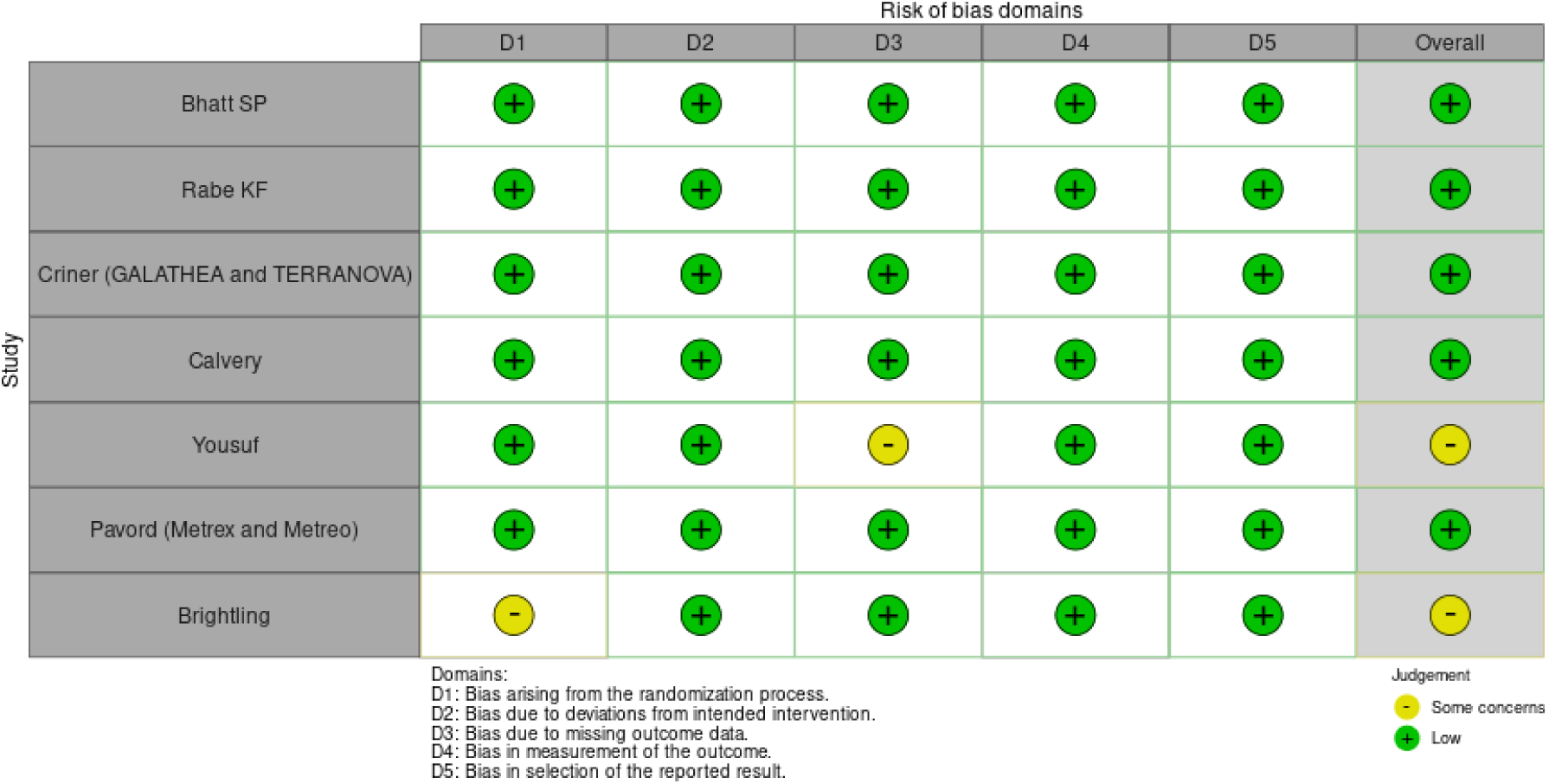

### Effects of interventions

All the included studies reported exacerbation rates. Seven studies, Yousuf (2023), Calverly (2017), Galathea (2019), Terranova (2019), Bhatt (2023), METREO (2017) and METREX (2017), measured quality of life using the St. George Respiratory Questionnaire. Additionally, six studies, Calverly (2017), Yousuf (2023), Galathea (2019), Terranova (2019), Bhatt (2023), Rabe KF (2021), reported on lung function.

Overall, there was substantial heterogeneity among most of the results. One possible explanation was the difference on the therapeutic targets (IL-5, IL-1R1, IL-33) and its impact on inflammation.

#### 1. Annualized exacerbation rate

The mean annual rate of exacerbations was reported in seven RCTs (18–20, 22–24). The use of biological therapy probably results in little to no effect on annual exacerbation rates compared with standard care (4152 patients; MD −0.12, 95% CI −0.23 to 0; I² = 33%). Moderate-certainty evidence supports the conclusion of no effect. (See GRADE pro 1.)

##### Analysis 1. Exacerbation rate

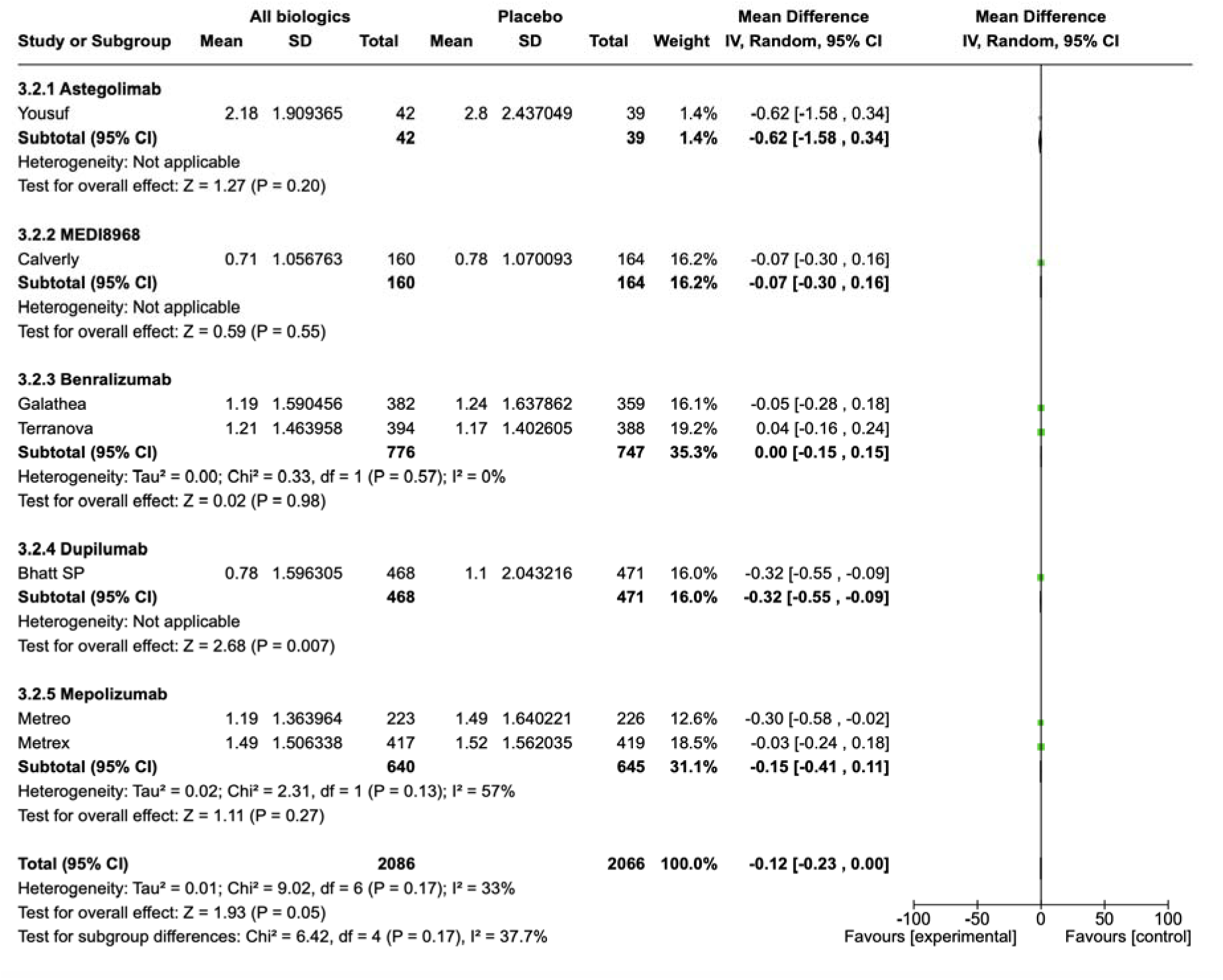

#### 2. Exacerbation Events in Patients

The exacerbation events were reported in four RCTs (18,19,24). The use of biological therapy probably results in little to no effect on on exacerbation events compared to the standard of care (2543 patients; RR 1.03, 95% CI 0.89 to 1.2; I2 = 73%). No differences were found by the type of biologic used. Moderate-certainty evidence supports the conclusion of no effect. (See GRADE pro 1.)

##### Analysis 2. Patients with exacerbations events

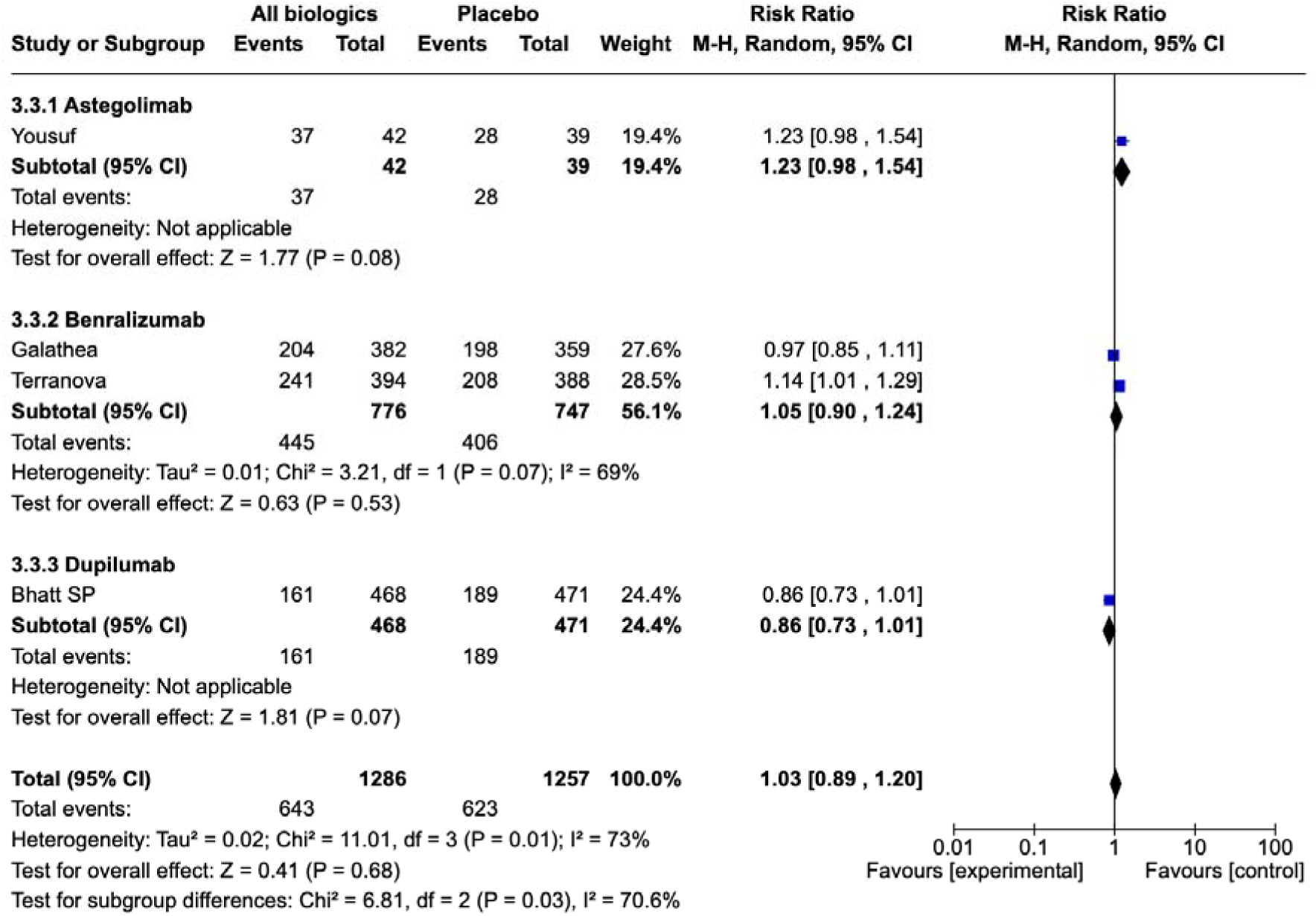

#### 3. Change in St. George Respiratory Questionnaire (SGRQ)

Change in SGRQ was reported in seven RCTs, (18–21). The evidence is very uncertain about the effect of the use of biological therapy on the quality of life measured by the SGRQ compared to the standard of care (3,906 patients; MD 0.05, 95% CI -0.04 to 0.14; I2 = 95,5%), therefore it is not possible to draw conclusions. Very low certainty evidence affected by imprecision and important inconsistency among studies’ results.

##### Analysis 3. SGRQ

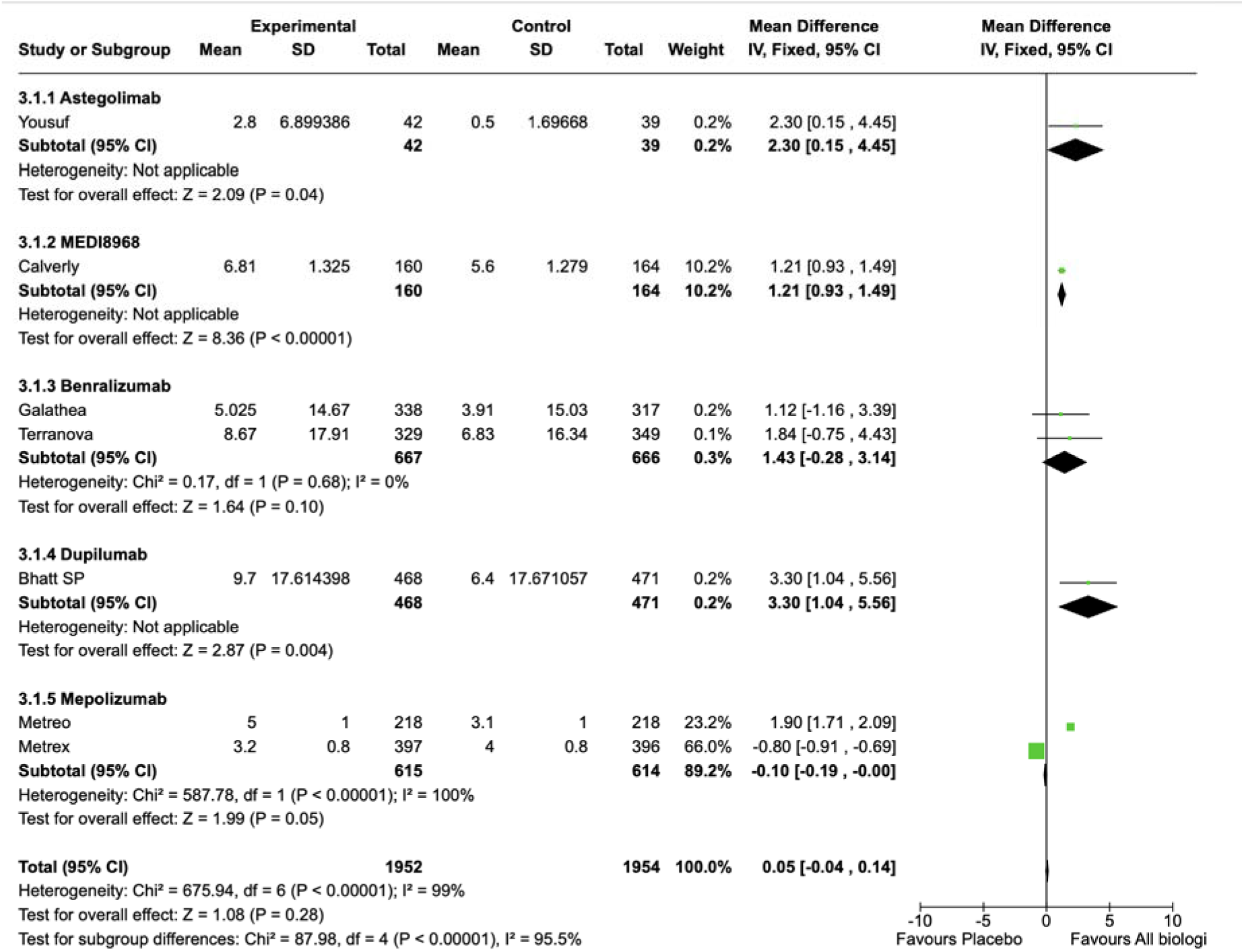

#### 4. Change in FEV1

Six randomized controlled trials (RCTs) reported the change in FEV1 (18–19, 21–23). The use of biological therapy may result in no differences in the FEV1 from baseline compared to the standard of care (2984 subjects; MD 0.02, 95% (CI) -0.02 to 0.06; I2 = 76.8%). Moderate-certainty evidence supports the conclusion of no effect.

##### Analysis 4. Change in FEV1

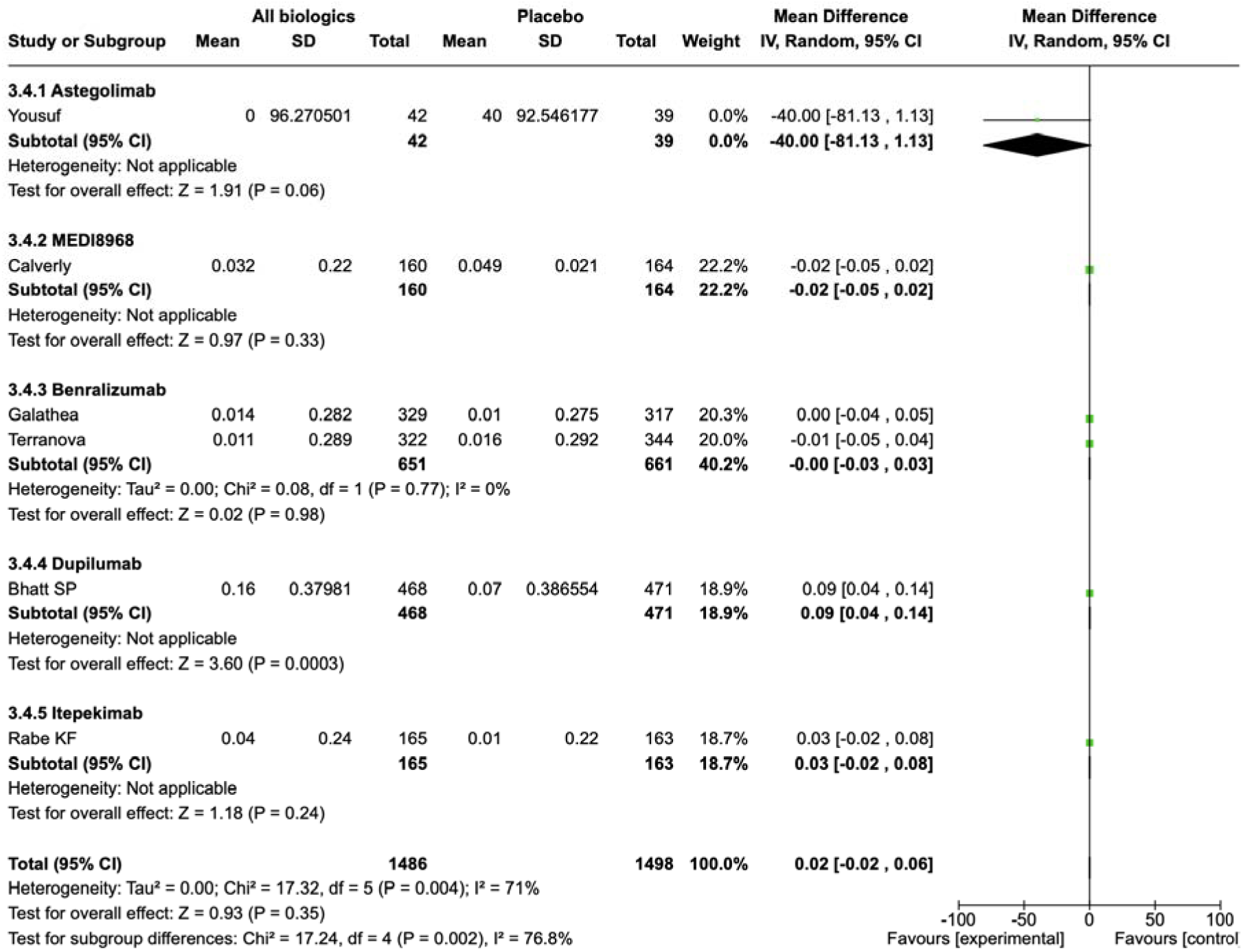

#### 5. Total adverse events

Eight randomized controlled trials (RCTs) reported the total number of adverse events (18–24). The use of biological therapy probably results in no difference in the total number of adverse events compared to the standard of care (4.041 subjects; RR 1.02; 95% CI 0.99 to 1.05; I2 = 0%). The evidence was rated as of high certainty for not effect.

##### Analysis 5. Adverse events (any) rate

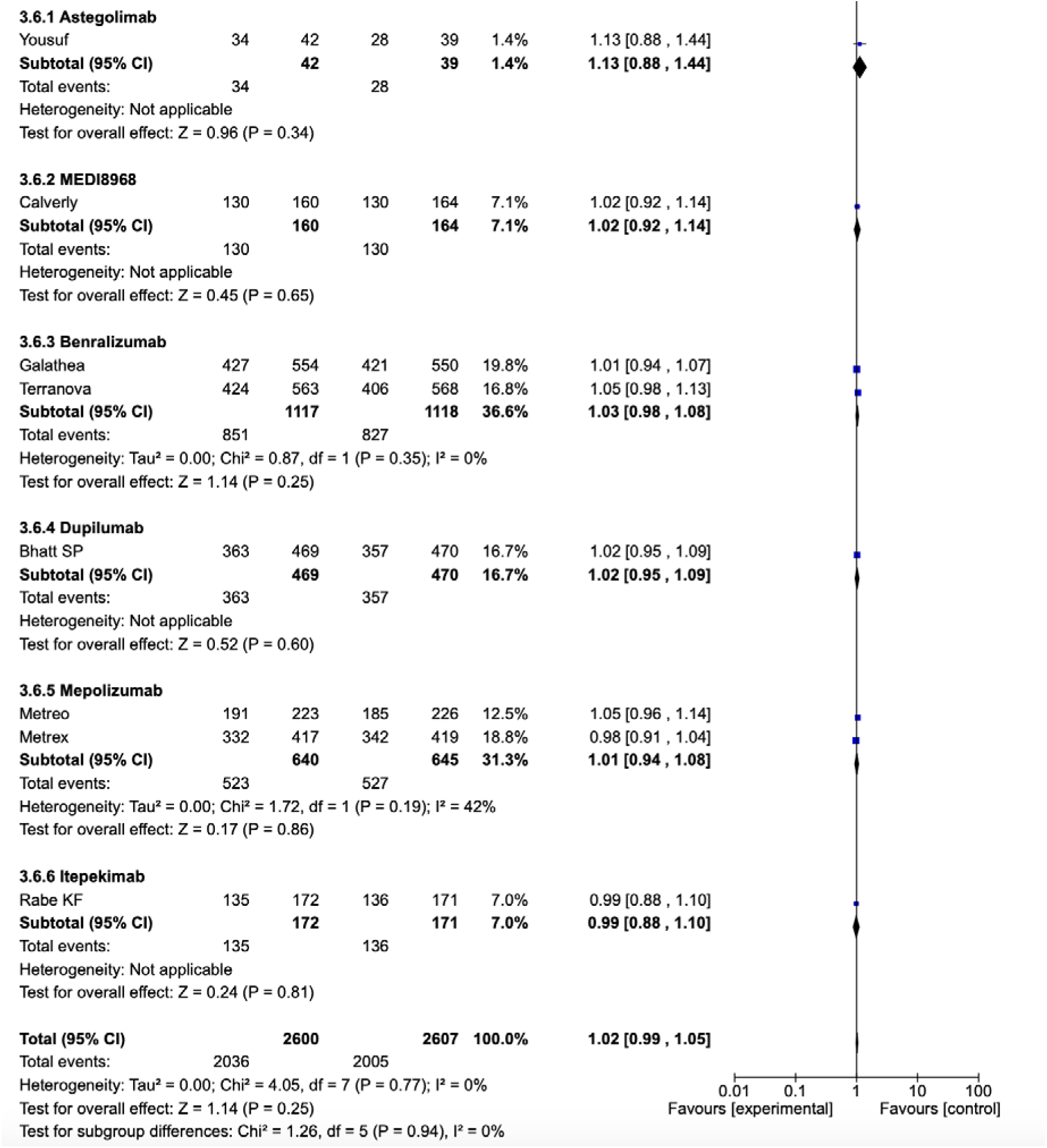

#### 6. Severe adverse events

Severe adverse events were reported in eight RCTs (18–24). Considering the MID cut off 0.75-1.25, the use of biological therapy may result in a small reduction to trivial or no effect in severe adverse events compared to the standard of care (5.207 subjects, (RR 0.84; 95% CI 0.77 to 0.93; I2 = 0%). The certainty of this outcome was moderate due to imprecision.

##### Analysis 6. Adverse events (severe)

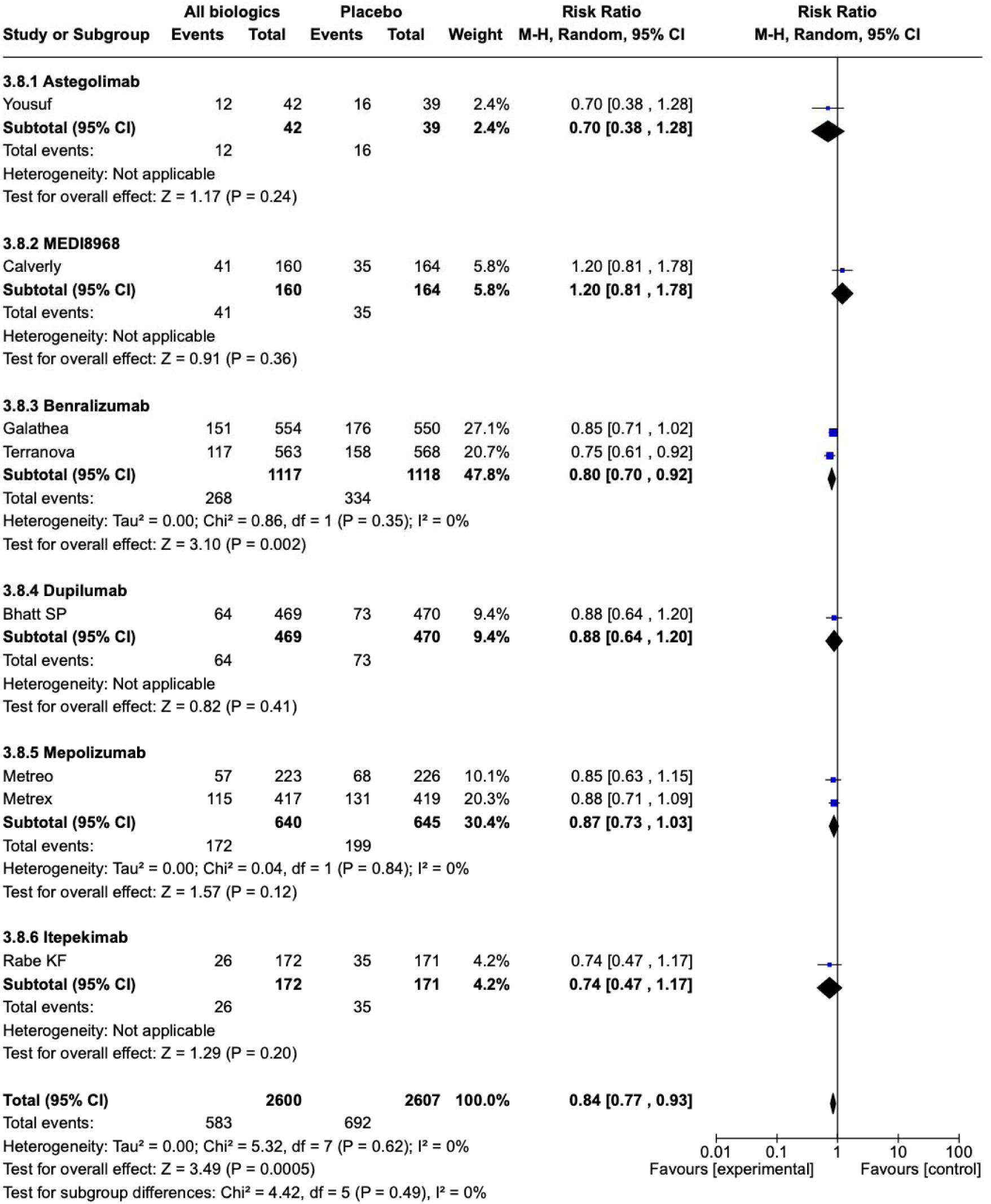

### Subgroup and sensitivity analysis

Given the inclusion of various molecules within the biological therapy, subgroup analyses were undertaken to assess their impact on outcomes.

### Effects of Mepolizumab on main outcomes

#### Quality of life

Change in SGRQ was reported in two studies (20). The use of biological therapy probably results in no difference in the St George questionnaire compared to standard of care (1.666 patients; MD 0.43, 95% CI -1.16 to 2.02; I2 = 100%). The certainty is very low due to inconsistency and serios impresicion.

#### Analysis 7. SGRQ

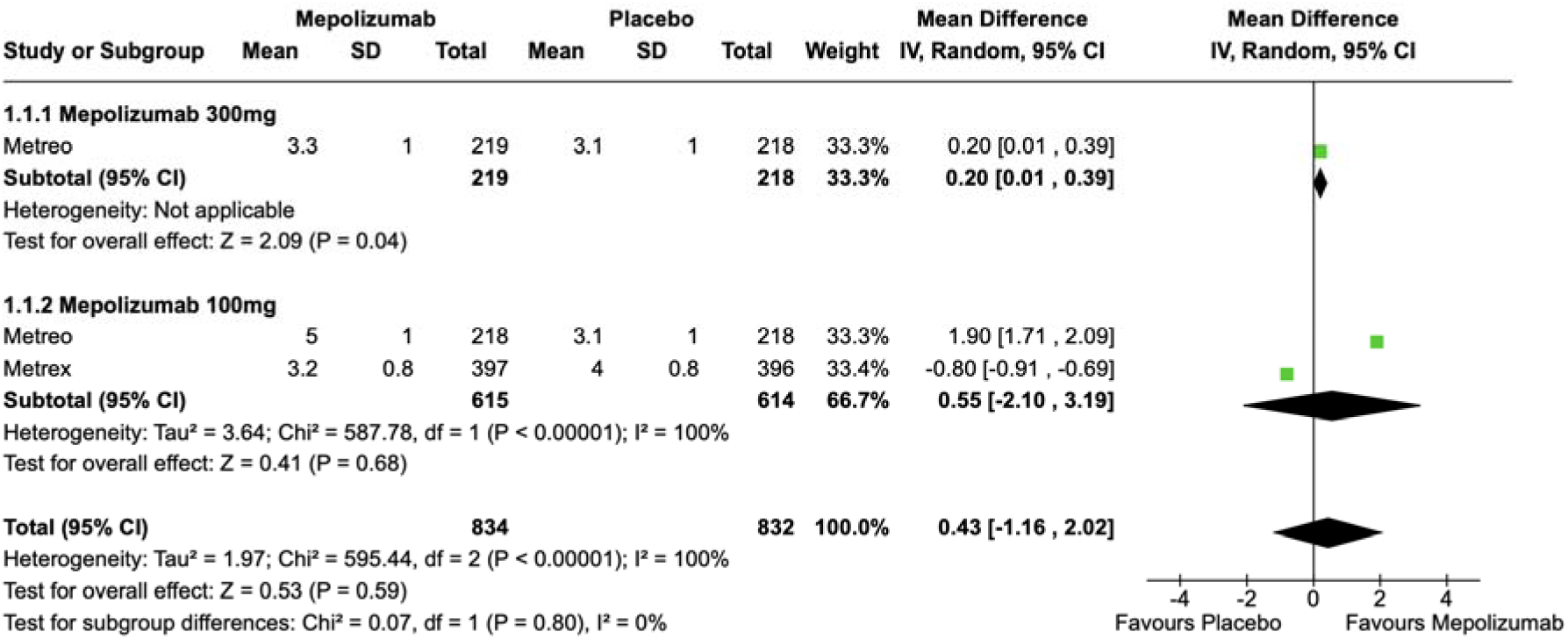

The responder’s rate in SGRQ was reported in two studies (20). The use of biological therapy results in no difference in the responder’s rate in the St George questionary score compared to standard of care. (1.343 patients; MD 1.11, 95% CI 0.97 to 1.27; I2 = 0%). The certainty of this outcome was very low

#### Analysis 1.2: SGRQ Responders (≥4-point improvement)

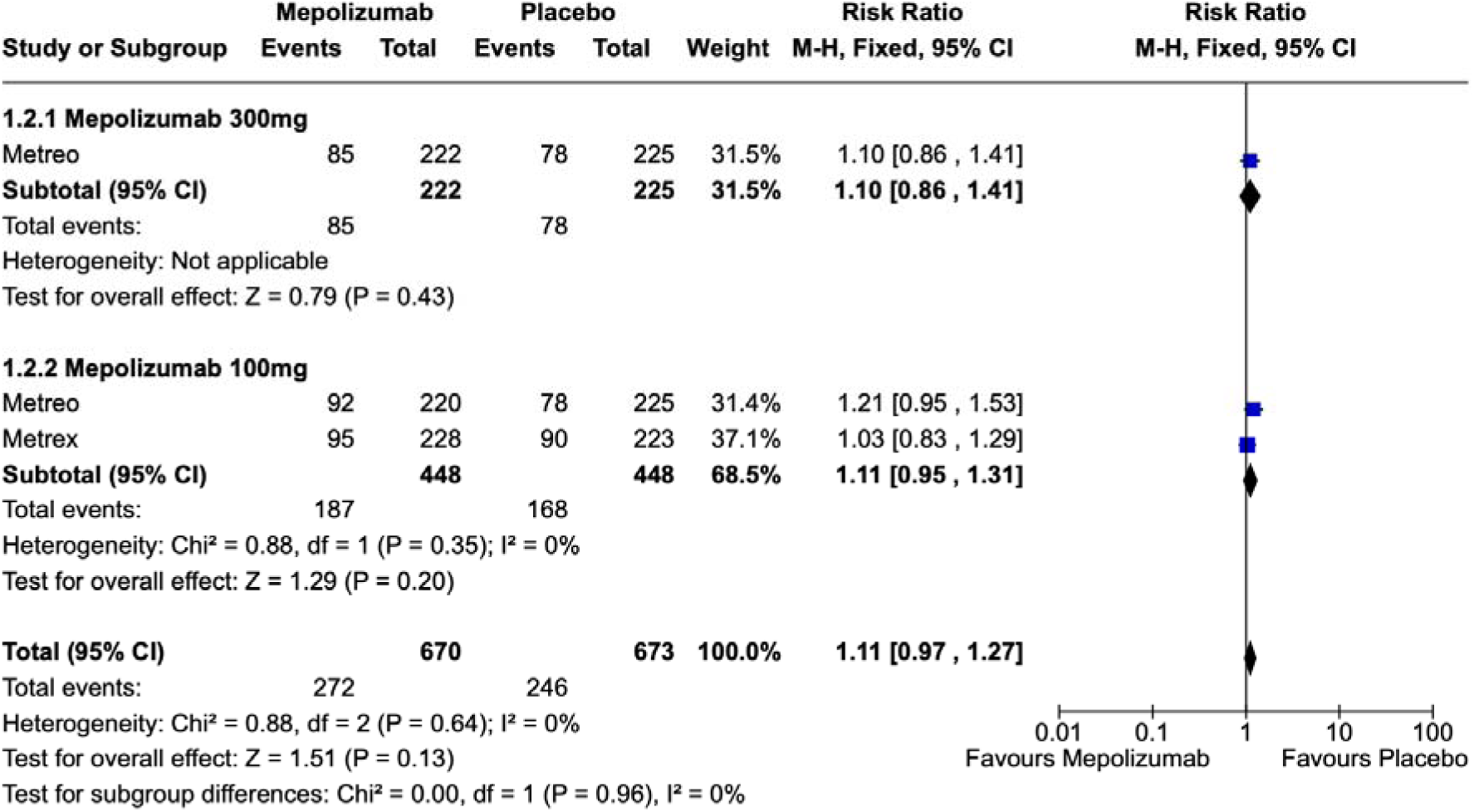

##### Exacerbations

The mean of exacerbation was reported in two RCTs (20). Biological therapy probably results in a small decrease to trivial or no effect in the exacerbation. (1.763 patients; MD - 0.15, 95% CI -0.29 to -0.01; I2 = 24%). The evidence is of moderate certainty because of imprecision.

#### Analysis 1.3: Mean of Exacerbation

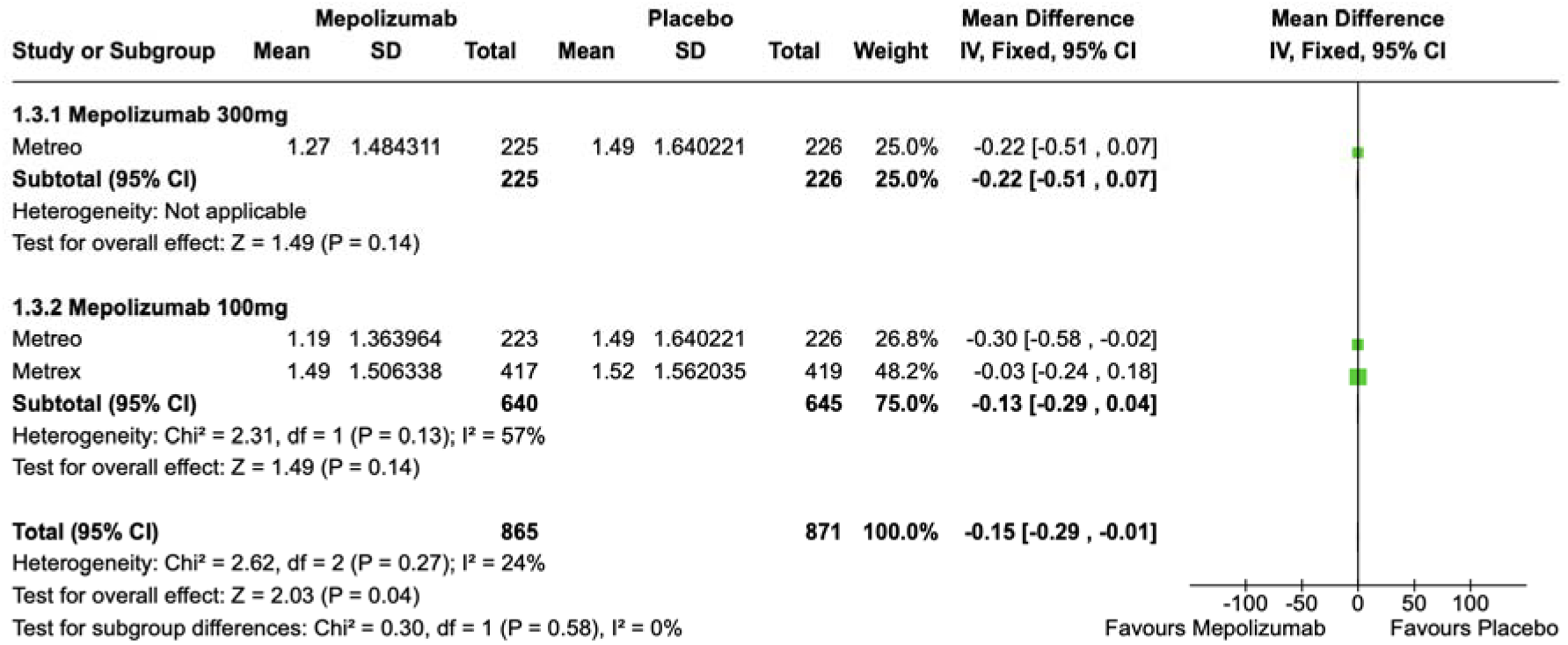

### Adverse Events

Two randomized controlled trials (RCTs) reported the total number of adverse events. The use of biological therapy results in no difference in the rate of adverse events compared to the standard of care (1.736 subjects; RR 1.02; 95% CI 0.97 to 1.08; I2 = 38%). The certainty of this outcome was high (evidence of no effect).

#### Analysis 1.4: Adverse Events (AEs) (any)

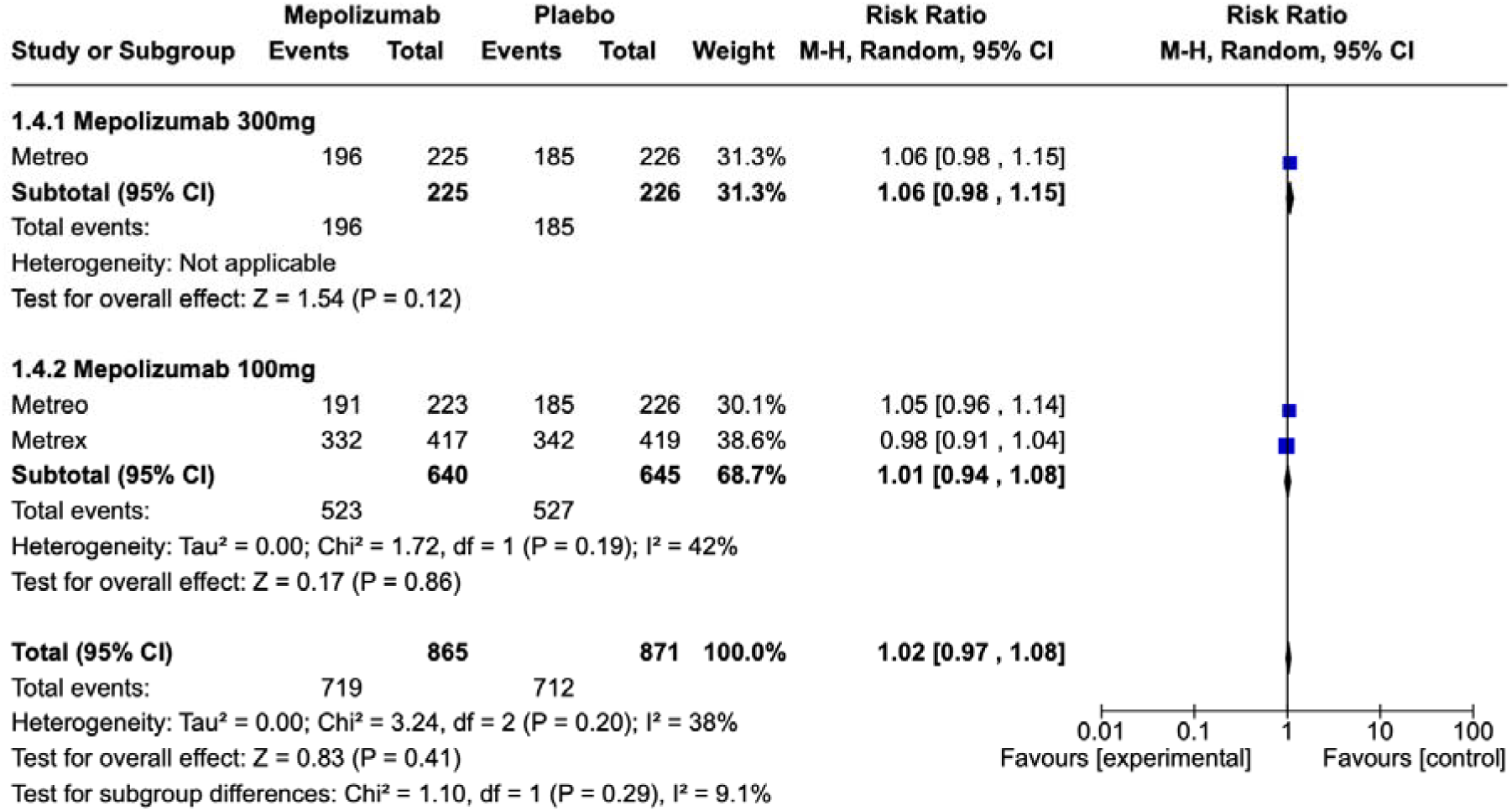

##### Severe adverse events

Two randomized controlled trials (RCTs) reported severe adverse events (20). The use of biological therapy results in no difference in the rate of severe adverse events compared to standard of care (1,736 subjects; RR 0.87, 95% CI 0.75 to 1.02; I² = 0%). The certainty of this outcome was high.

#### Analysis 1.5: Severe Adverse Events

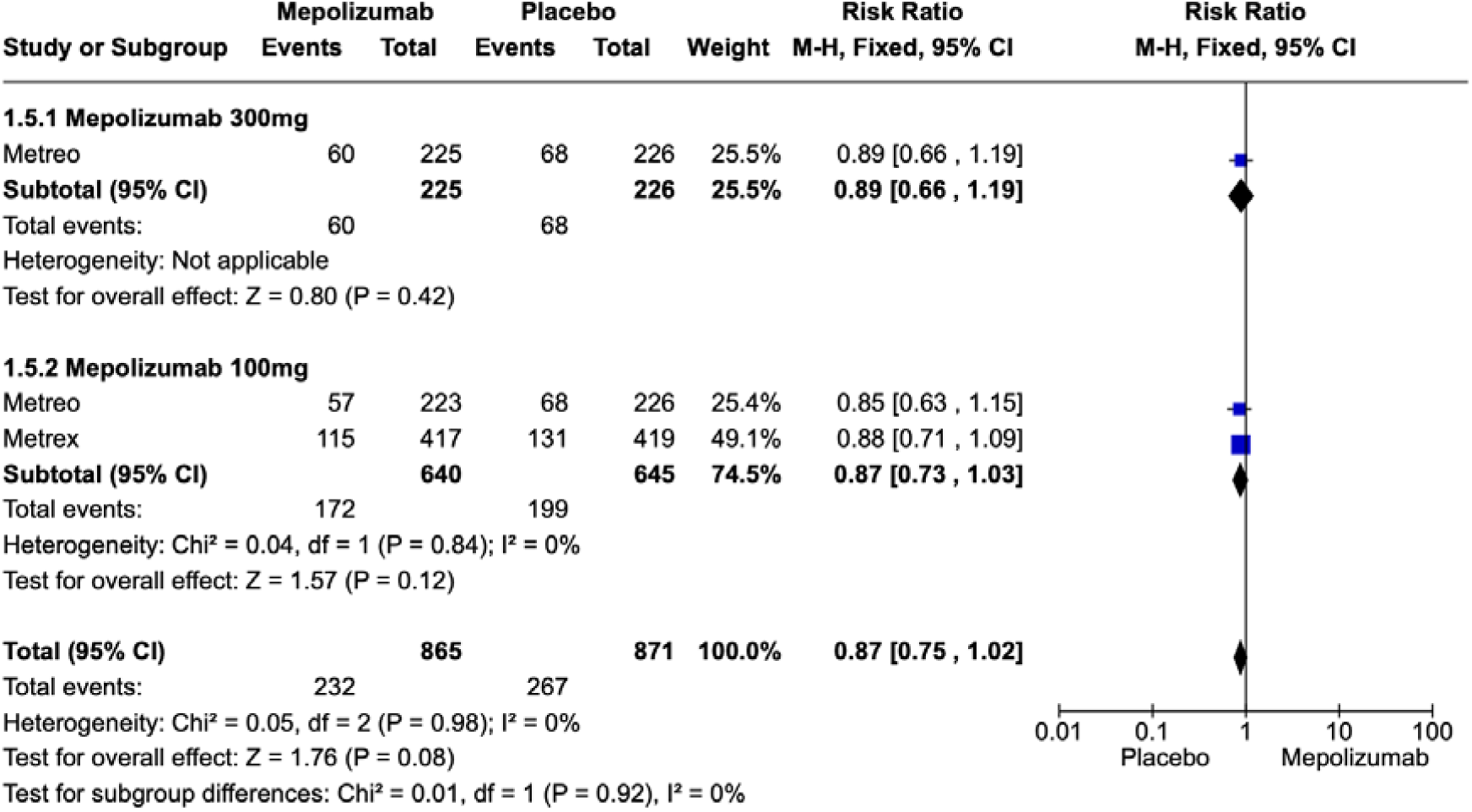

### Benralizumab

#### Quality of life

Change in SGRQ was reported in three studies (23,24), The use of Benralizumab results in no difference in the St George questionary score (3.465 patients; MD 1.33 95% CI 0.29 to 2.37; I2 = 0%). The certainty of this outcome was high

##### Analysis 2.1: SGRQ

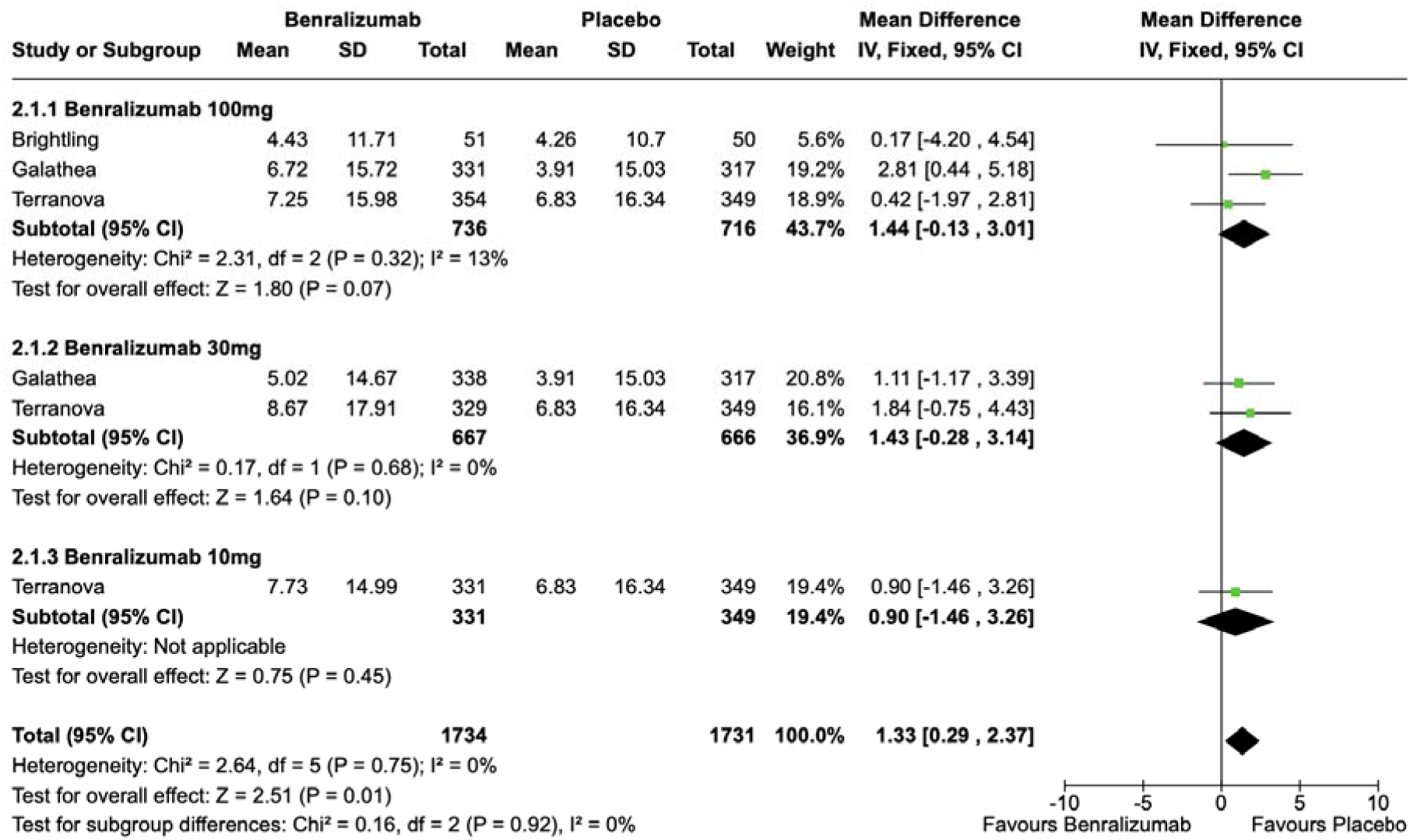

#### Exacerbations

The mean of exacerbation was reported in three RCTs (23,24). Benralizumab results in a small decrease in the mean of exacerbations compared to standard of care. (3.903 patients; MD -0.09, 95% CI -0.18 to 0; I2 = 16%). The certainty of evidence is high

##### Analysis 2.2: Mean of Exacerbation

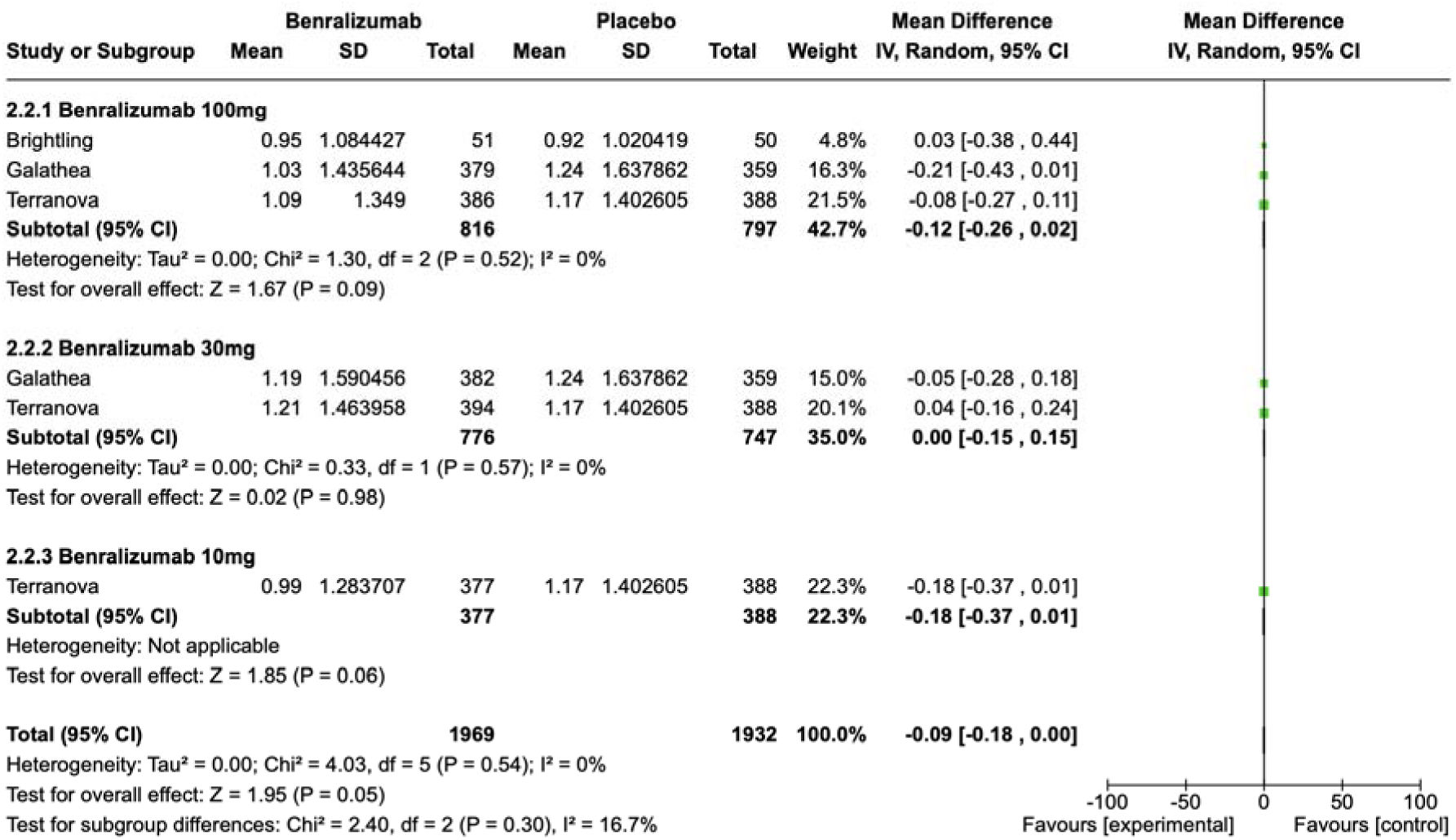

#### Patients with exacerbations

The patient with present exacerbation was reported in two RCTs (24). The use of Benralizumab probably results in no difference in the number of patients with exacerbations compared to the use of standard of care. (3.800 patients; RR 1.02, 95% CI 0.97 to 1.09; I2 = 10%). The evidence is affected mainly by imprecision.

##### Analysis 2.3: Patients with exacerbations

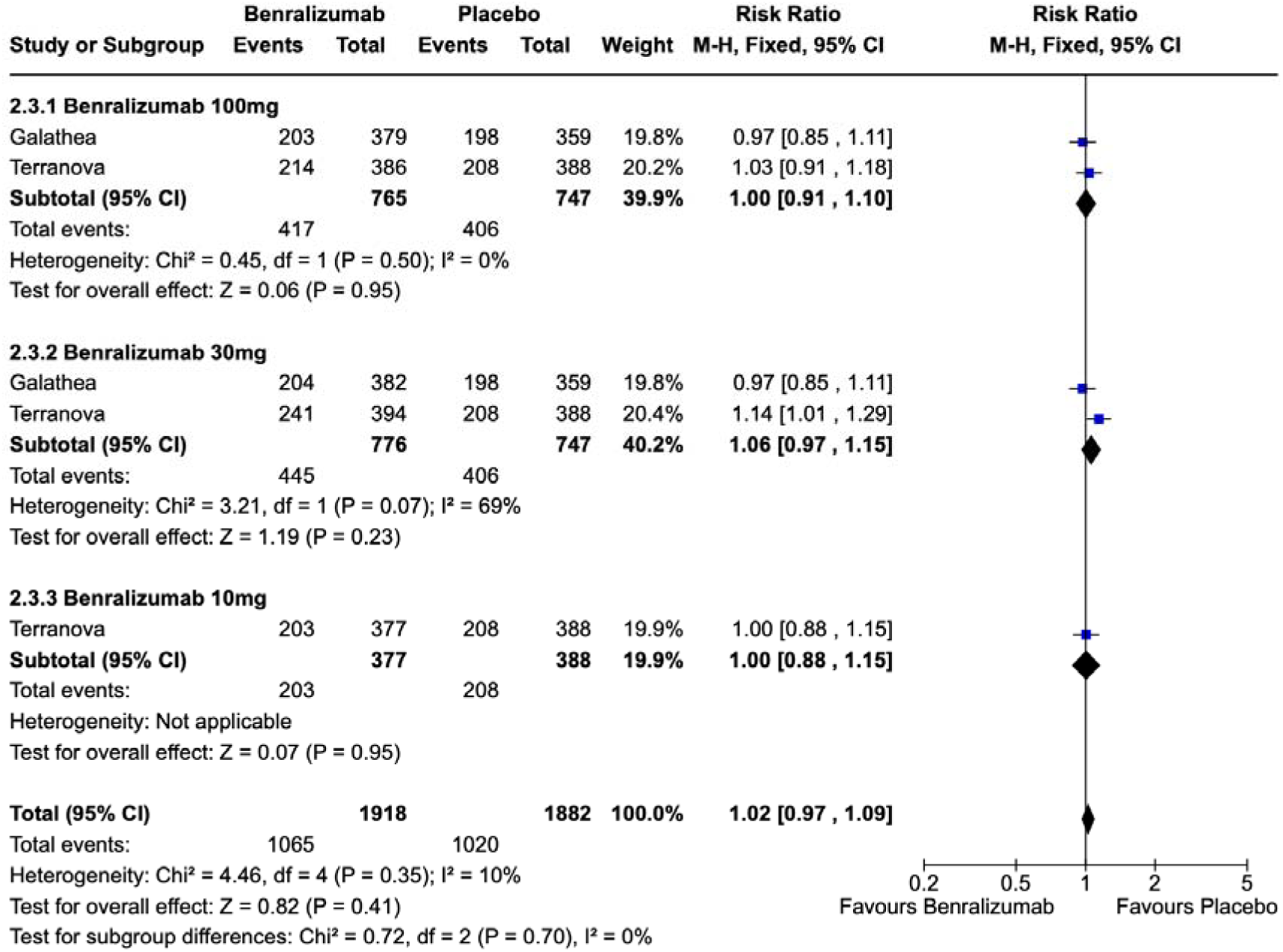

#### Lung function

Change in FEV1 was reported in three studies (23,24). The use of Benralizumab probably results in no difference in lung function compared to the use of standard of care (3.393 patients; MD 0.01 95% CI -0.01 to 0.03; I2 = 0%). The certainty of evidence is moderate, mainly affected by imprecision.

##### Analysis 2.4: Change in FEV1

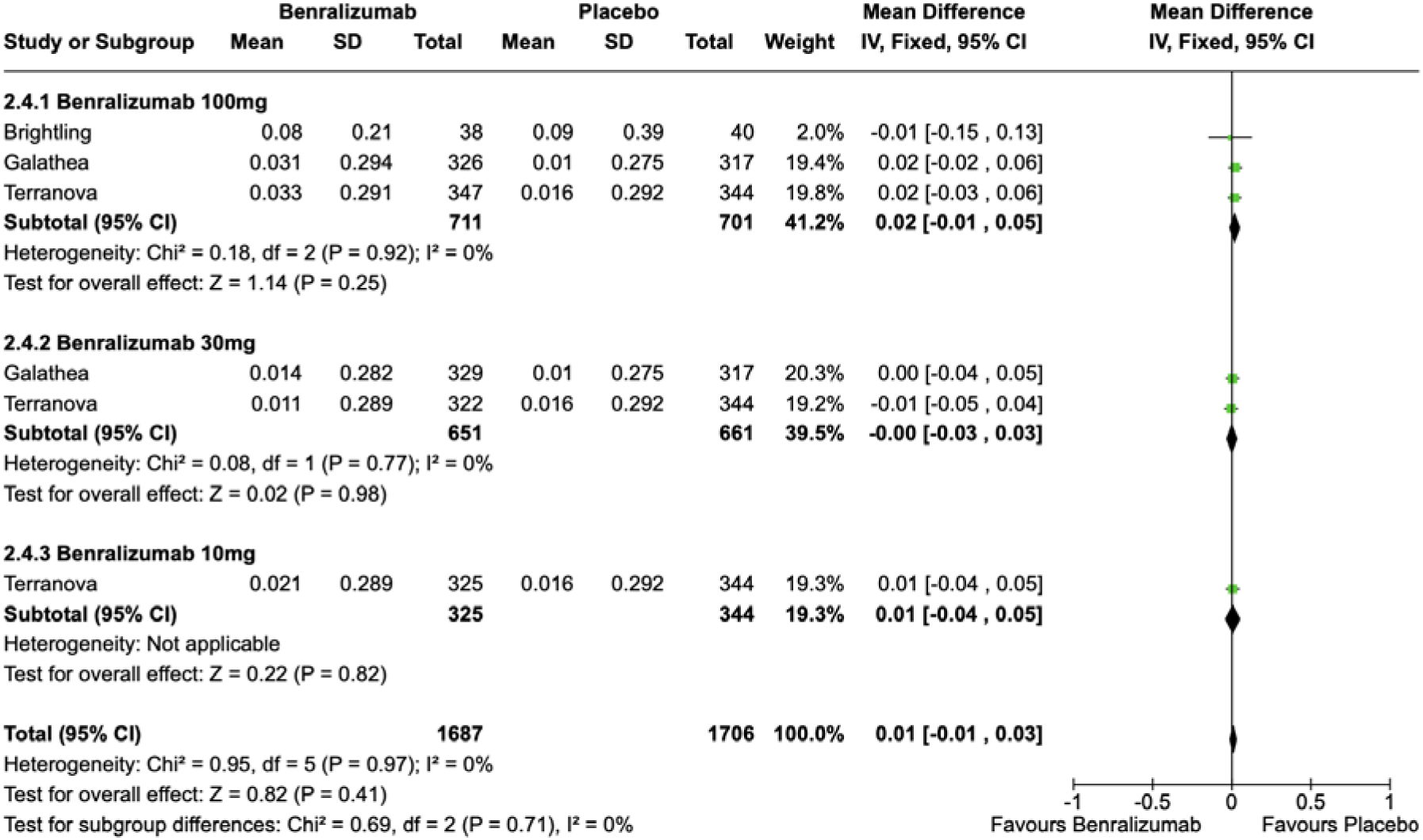

#### Adverse Events

Three randomized controlled trials (23,24) reported any adverse events. The use of benralizumab results in no difference in adverse events compared to standard of care. (5.682 subjects; RR 1.02, 95% CI 0.99 to 1.05; I² = 0%). The certainty of evidence is high.

##### Analysis 2.5: Adverse Events (any)

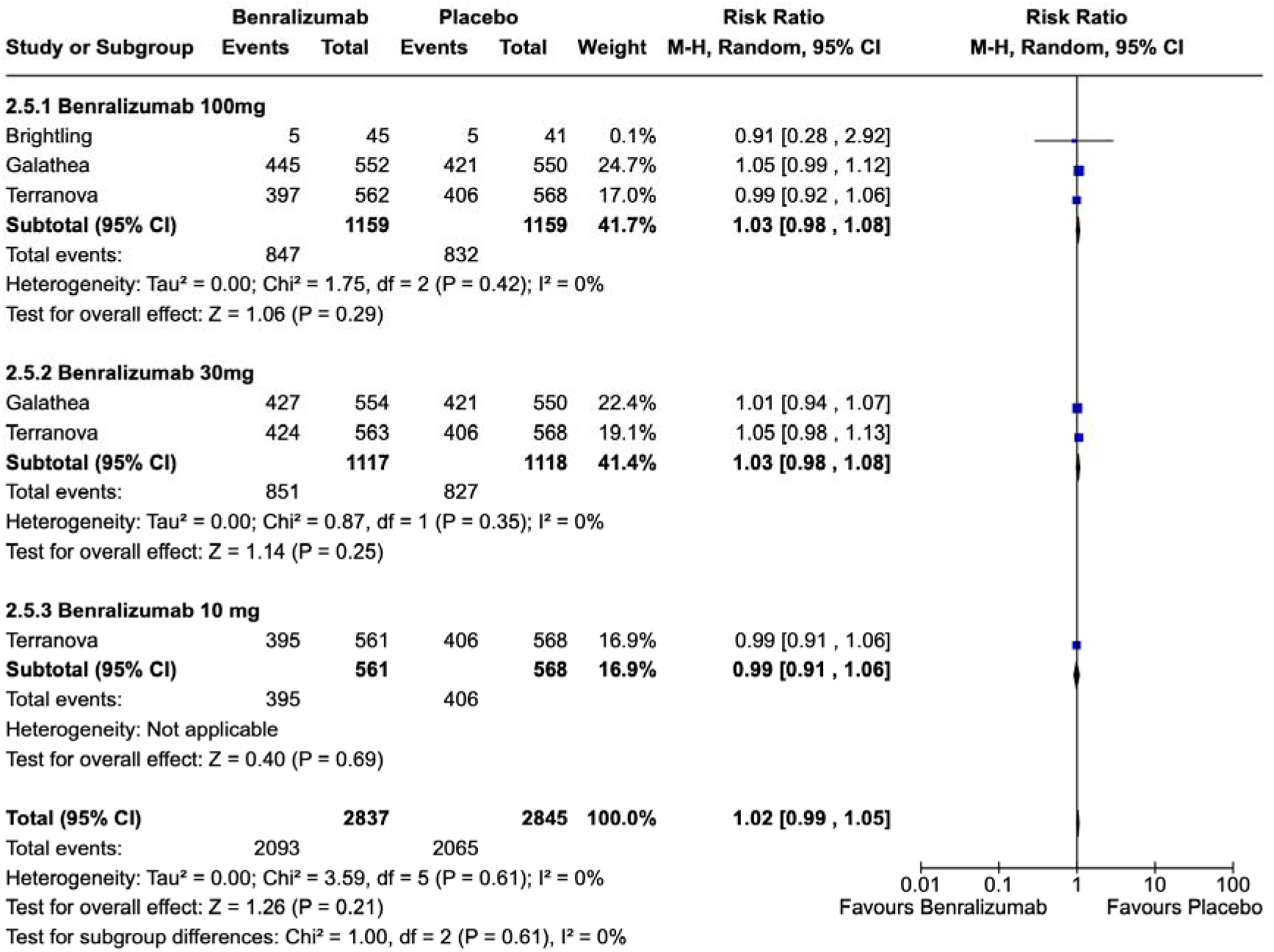

#### Severe adverse events

Three randomized controlled trials (RCTs) reported severe adverse events (23,24). Benralizumab results in a decrease in the rate of severe adverse events compared to the use of standard of care. (5.682 subjects; RR 0.87, 95% CI 0.79 to 0.96; I² = 27%)

##### Analysis 2.6: Severe Adverse Events

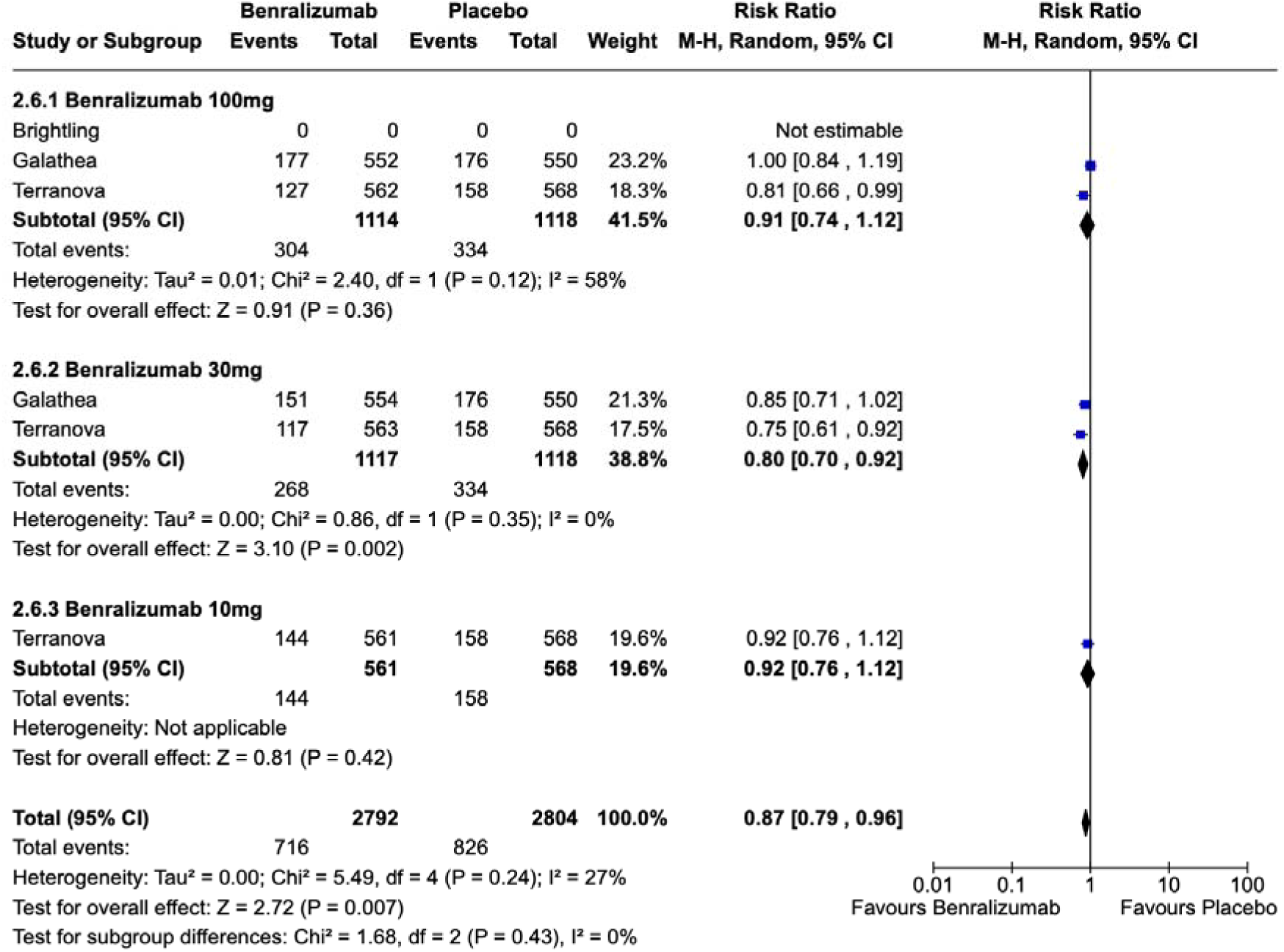

## Discussion

### Overview of findings

This baseline review, part of the living evidence synthesis, included nine randomized controlled trials (RCTs) that evaluated the use of biological therapy versus placebo in patients with COPD. Three studies compared benralizumab to placebo (Brightling 2014, GALATHEA 2019, and TERRANOVA 2019), two evaluated mepolizumab versus placebo (METREO 2017 and METREX 2017), one compared MEDI8968 to placebo (Calverley 2017), Yousuf 2022 compared astegolimab with placebo, Bhatt 2023 evaluated dupilumab against placebo, and Rabe 2021 compared itepekimab to placebo.

Current evidence suggests that biological therapies for COPD provide little to no clinical benefit over standard care in terms of exacerbation rates, lung function, or safety, with uncertain effects on quality of life. Specifically: Biological therapy probably has little to no effect on annual exacerbations compared with standard care. There is likely no meaningful difference in lung function (FEV1) compared with standard care. Biologicals probably result in no difference in the number of total adverse events.

Regarding the effect on quality of life, measured by the SGRQ compared with standard care, the evidence is very uncertain; therefore, no conclusions can be drawn.

Overall, the certainty of the evidence supporting these statements is generally moderate, but is particularly limited by imprecision in estimates when considering thresholds for the minimal important difference (MID).

### Overall completeness and applicability of evidence

Evidence regarding the impact of biologics in the annualized exacerbation rates was available in all studies. However, the information regarding lung function is limited, particularly in the absence of data on mepolizumab and its influence on FEV1. Additionally, the research conducted by Rabe KF et al. (pertaining to Itepekimab) failed to document any impact on quality of life, as assessed by the St. George questionnaire.

Furthermore, comprehensive Phase 3 studies are still pending regarding MEDI8968, Itepekimab and Astegolimab.

In summary, while the analysis sheds light on the potential of biological therapies for COPD management, the current evidence base remains inconclusive in its impact in lung function and quality of life, highlighting the need for further research and standardization in study design and participant selection criteria.

### Comparison to other similar publications

In recent years, research in this field has found a potential reduction in the rate of moderate to severe exacerbations with anti-IL5 use. A Cochrane review (12) reported a decrease in the rates of moderate to severe exacerbations with the use of mepolizumab. This reduction was greater in those patients with higher eosinophil counts (defined as ≥ 150 cells per mm³ at screening or ≥ 300 cells per mm³ in the year before trial entry); however, no improvement was found for other outcomes, such as the impact on quality of life as measured by the St. George’s score and impact on lung function. On the other hand, Ohnishi’s (13) article showed similar results, with a reduction in the rate of moderate to severe exacerbations with the use of mepolizumab. Additionally, an integrated analysis showed a reduction in exacerbations leading to emergency department visits or hospitalizations with the use of Benralizumab. There was no improvement in the SGRQ score.

### Limitations and strengths

This overview of systematic reviews has multiple strengths. We developed this review using a robust predefined methodology stated in a registered protocol.

All steps were done in duplicate thus limiting errors throughout the review process. In addition, the evidence was summarized using GRADE methodology which is a highly transparent methodology for evidence presentation and development. We used high and moderate quality systematic reviews based on AMSTAR tool.

### Clinical implications

This review is a milestone in creating evidence-based guidance regarding biological therapy as a COPD treatment. With the current evidence, it is not possible to recommend the use of biologic therapy for the treatment of chronic obstructive disease. However, in this review we found results suggesting that the use of some biologics could be used in a selected group of patients with COPD and eosinophilia. More evidence is needed in this group of patients.

### Implications for research

This review showed a lack of high-quality systematic reviews with biologic therapy in patients with COPD and type 2 inflammation.

This evidence synthesis is part of a larger project set up to put the living evidence approach into practice. This project aims to produce multiple parallel living systematic reviews relevant to inform decisions, following the higher standards of quality in evidence synthesis production. We believe that our methods are well suited to handle the evidence that is to come, including evidence on the role of biological therapy in COPD.

We have identified several ongoing studies addressing this question, including 2 randomized trials, which will provide valuable evidence to inform researchers and decision makers soon.

During the next year, we will maintain a living, web-based, openly available version of this review at the Living Evidence to Inform Health Decisions website (https://livingevidenceframework.com/en/lesr/), and we will re-submit the review for publication every time the conclusions change or whenever there are substantial updates. Our systematic review aims to provide a high-quality, up-to-date synthesis of the evidence that is useful for clinicians and other decision makers.

## Data Availability

All data produced in the present study are available upon reasonable request to the authors

https://osf.io/jtw35/overview

## Acknowledgments

We would like to acknowledge the contribution of the Living Evidence to Inform Health Decisions Program research group and the methodologic team of Epistemonikos Foundation to the development of this review.Roles and contributions

García O. and Rojas M.X. conceptualized the study and provided overall supervision of the project. Rojas M.X. and Urrutia G. contributed methodological advice and support. Screening and selection were performed by Poloni D., Torres-Lopez L., and Díaz L. Data extraction and risk of bias assessment were carried out by Torres-Lopez L. and Díaz L. The meta-analyses were conducted by Auladell A. Evidence synthesis was undertaken by García O., Rojas M.X., and Díaz L. The original draft of the manuscript was prepared by García O., Díaz L., and Rojas M.X., all authors contributed to writing, review, and editing. All authors approved the final version of the manuscript.

## Funding

This project has been developed as part of the Living Evidence to Inform Health Decisions (LE-IHD) program’s project: “Strengthening decision-making capacity in the Spanish Health System through living evidence: An innovative framework” funded by Instituto Carlos III (ISC-III) Grant PI21/01564; Madrid, Spain and co-funded by and co-funded by the European Union through FEDER and the Horizon’s 2020 research and innovation programme (Grant Agreement MSCA-IF-EF-ST #894990), awarded to María Ximena Rojas for the development of the Living Evidence to Inform Health Decisions project.

The LE-IHD program provided training, support, and tools at no cost for supporting the development of this project.

There is no institutional funding.

## Protocol OSF registration

CRD42020181032

## Ethics

As researchers will not access information that could lead to the identification of an individual participant, obtaining ethical approval was waived.

## Appendix 1. Search strategy

The main search source will be Epistemonikos database (https://www.epistemonikos.org), a comprehensive database of systematic reviews and other types of evidence, maintained by screening multiple information sources to identify systematic reviews and their included primary studies, including Cochrane Database of Systematic Reviews, MEDLINE, EMBASE, CINAHL, PsycINFO, LILACS, DARE, HTA Database, Campbell database, JBI Database of Systematic Reviews and Implementation Reports, EPPI-Centre Evidence Library (6).

For this project, an additional search was performed on MEDLINE in order to identify randomized trials/primary studies not included in reviews.

### Boolean search strategy

|  |  |
| --- | --- |
| **Source** (*e.g., bibliographic databases, trial registries, web-based repositories*) |  |
| **Boolean Strategy** (*terms for population, intervention, boolean connectors*) | (((chronic* AND obstructive* AND (pulmonary* OR lung* OR airway*) AND disease*) OR emphysema* OR "obstructive lung disease" OR "obstructive lung diseases" OR (chronic* AND airflow* AND (obstruction* OR limitation*)) OR (hypercapni* AND respiratory* AND failure*) OR aecopd OR copd) AND (targeted* OR biologic* OR monoclonal* OR omalizumab* OR Xolair* OR mepolizumab* OR Nucala* OR reslizumab* OR Cinqair* OR Cinqaero* OR benralizumab* OR Fasenra* OR dupilumab* OR Dupixent* OR Lebrikizumab* OR MILR1444A* OR RG3637* OR TNX650* OR "RG-3637" OR "RG 3637" OR "TNX-650" OR "TNX 650" OR tralokinumab* OR Adtralza* OR Adbry* OR CAT354* OR "CAT-354" OR "CAT 354" OR tezepelumab* OR Tezspire* OR MEDI9929* OR |
|  | AMG157* OR "AMG-157" OR "AMG 157" OR brodalumab* OR Siliq* OR Kyntheum* OR KHK4827* OR "KHK-4827" OR "KHK 4827" OR AMG827* OR "AMG-827" OR "AMG 827" OR ligelizumab* OR QGE031* OR "QGE-031" OR "QGE 031" OR astegolimab* OR mstt1041a* OR "mstt-1041a" OR "mstt 1041a" OR "mstt1041-a" OR "mstt-1041-a" OR "mstt 1041-a" OR rg6149* OR "rg-6149" OR "rg 6149" OR itepekimab* OR tozorakimab*) AND ("critical review" OR "electronic search" OR "evidence-based analysis" OR "evidence-based review" OR "literature search" OR "meta analysis" OR "meta synthesis" OR "meta-analyse" OR "meta-analytic review" OR "meta-study" OR "meta-synthesis" OR "metaanalysis" OR "metasynthesis" OR "meta-analysis" OR "pooled effect" OR "random-effects model" OR "systematic quantitative review" OR "systematically searched" OR "systemic review" OR (review AND randomized) OR (systematic AND review) OR MEDLINE OR "literature review" OR PubMed)) |
| **Type of studies** (*e.g. RCTs, NRS, case-control, cohorts*) | RS, RCT |
| **Date of coverage** (*from dd/mm/yyyy*) | 30/03/2023 |
| **Filters (if any)** | language: english and spanish |

## Appendix 2. Characteristics of studies.

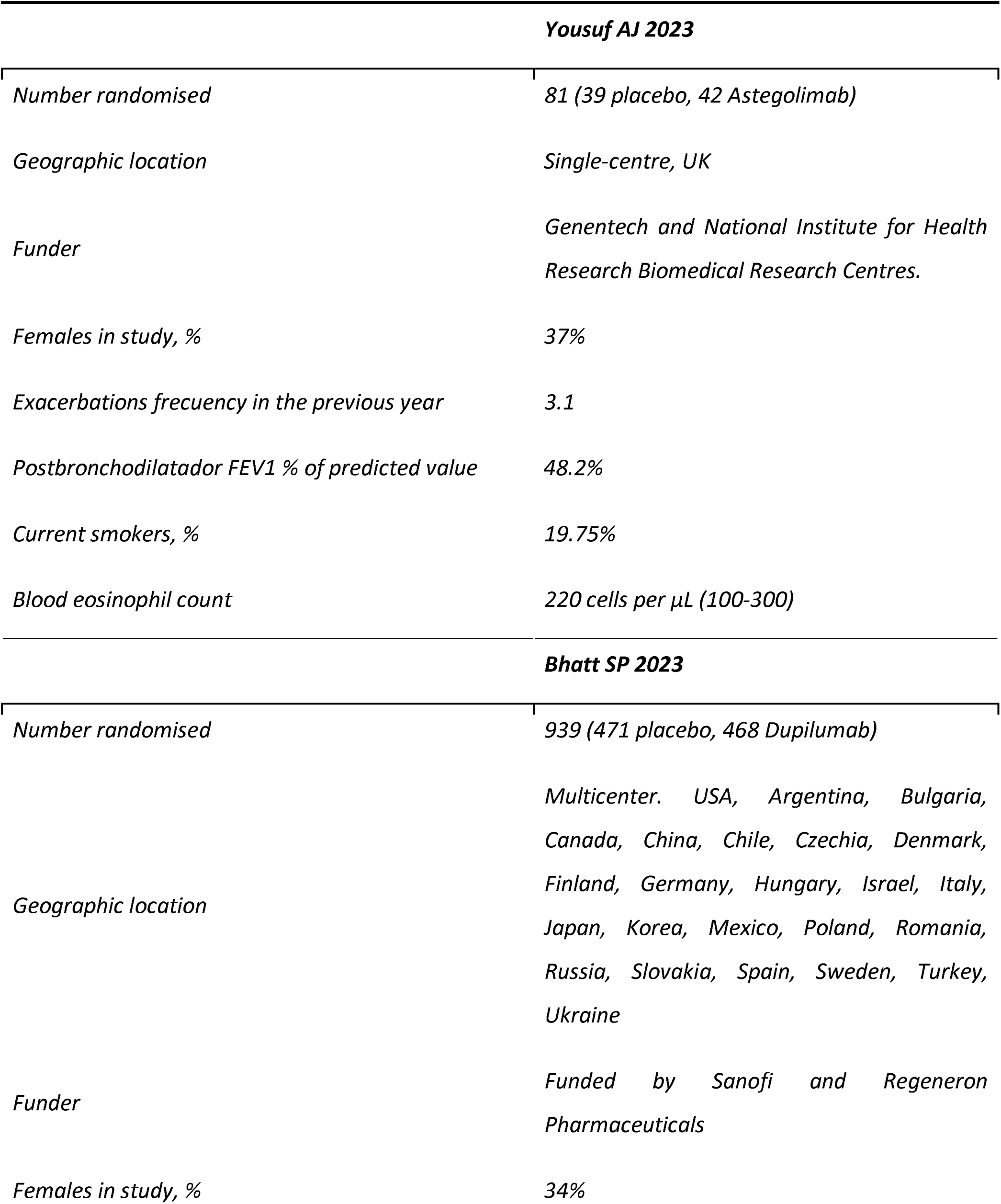
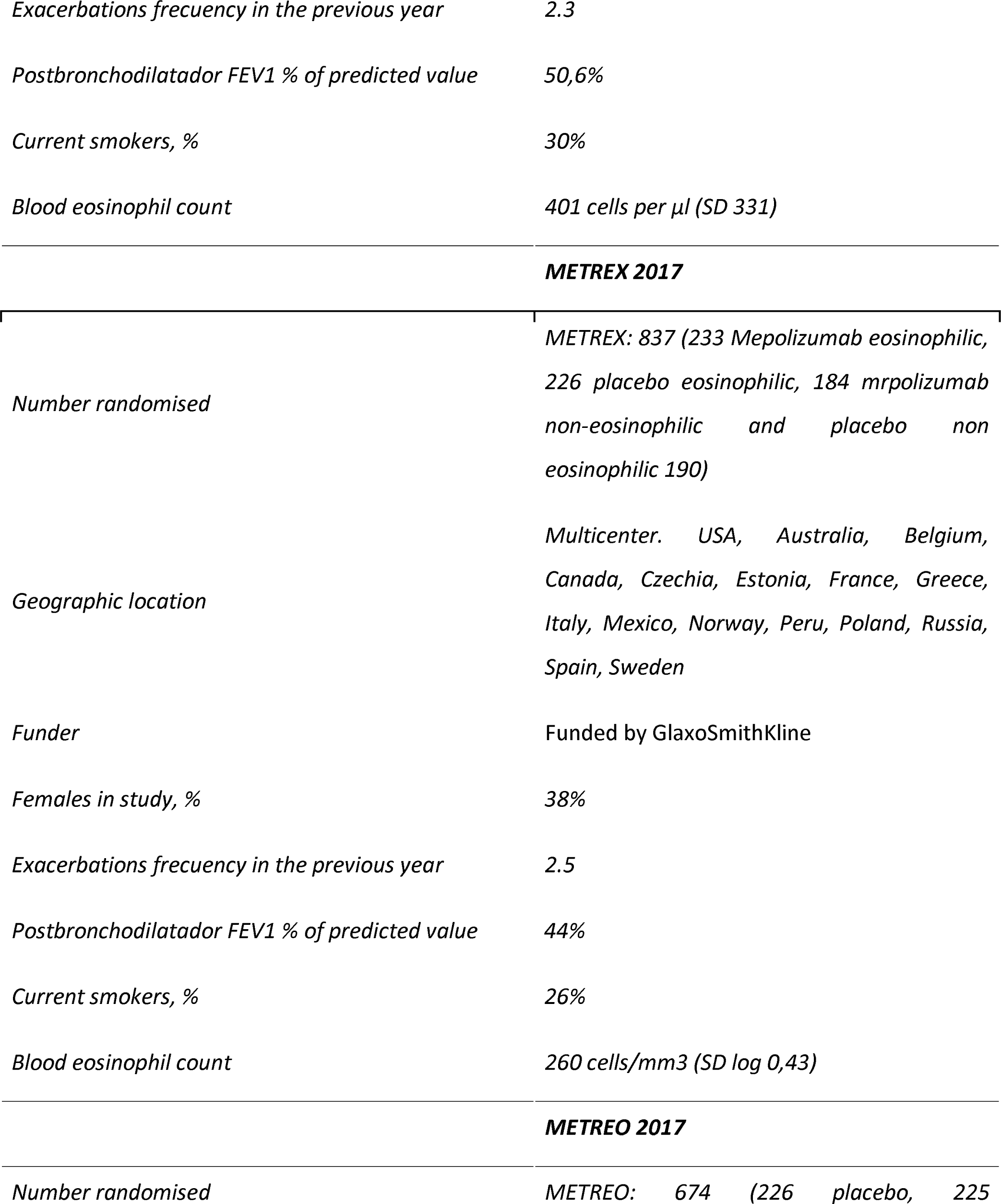
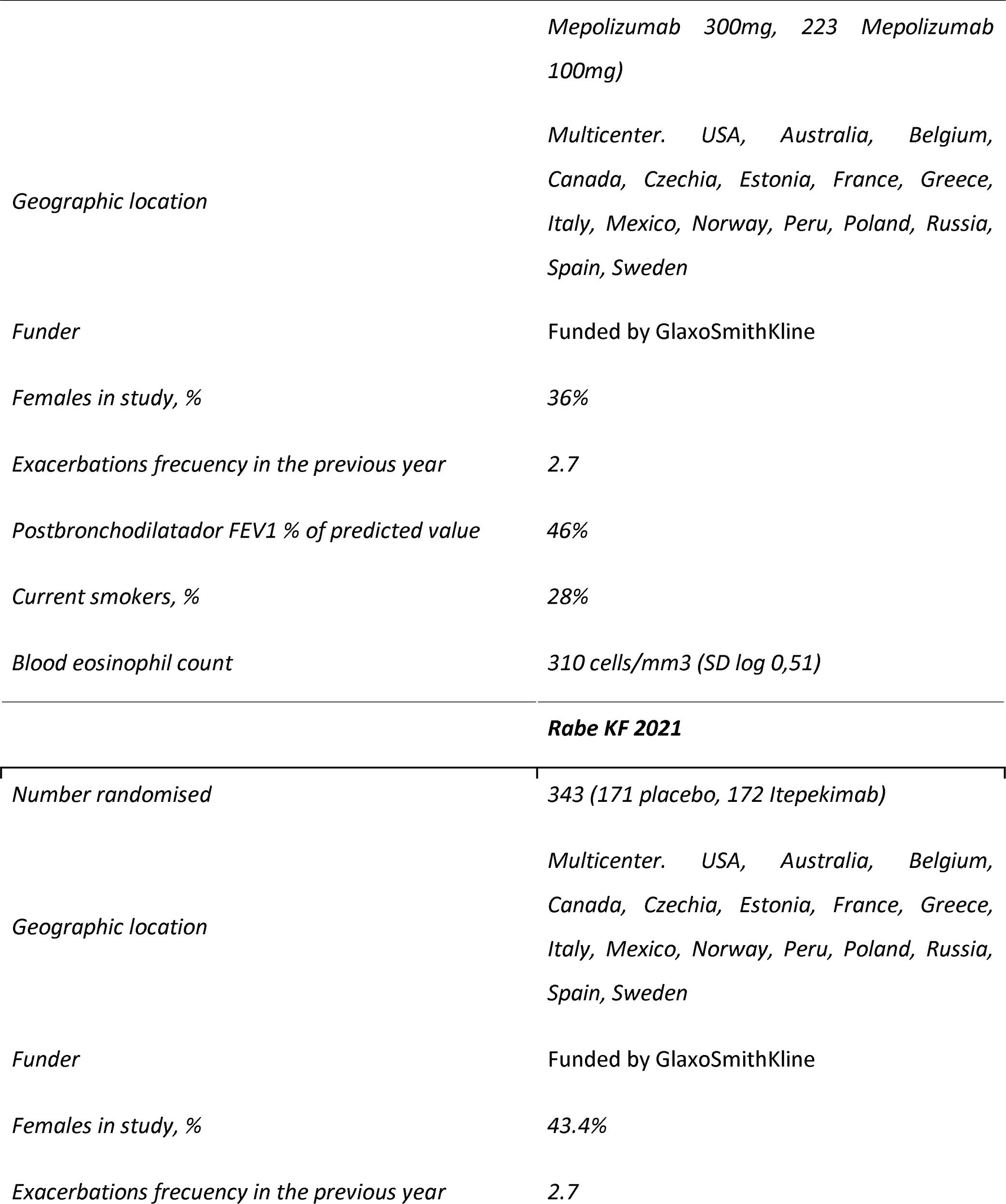
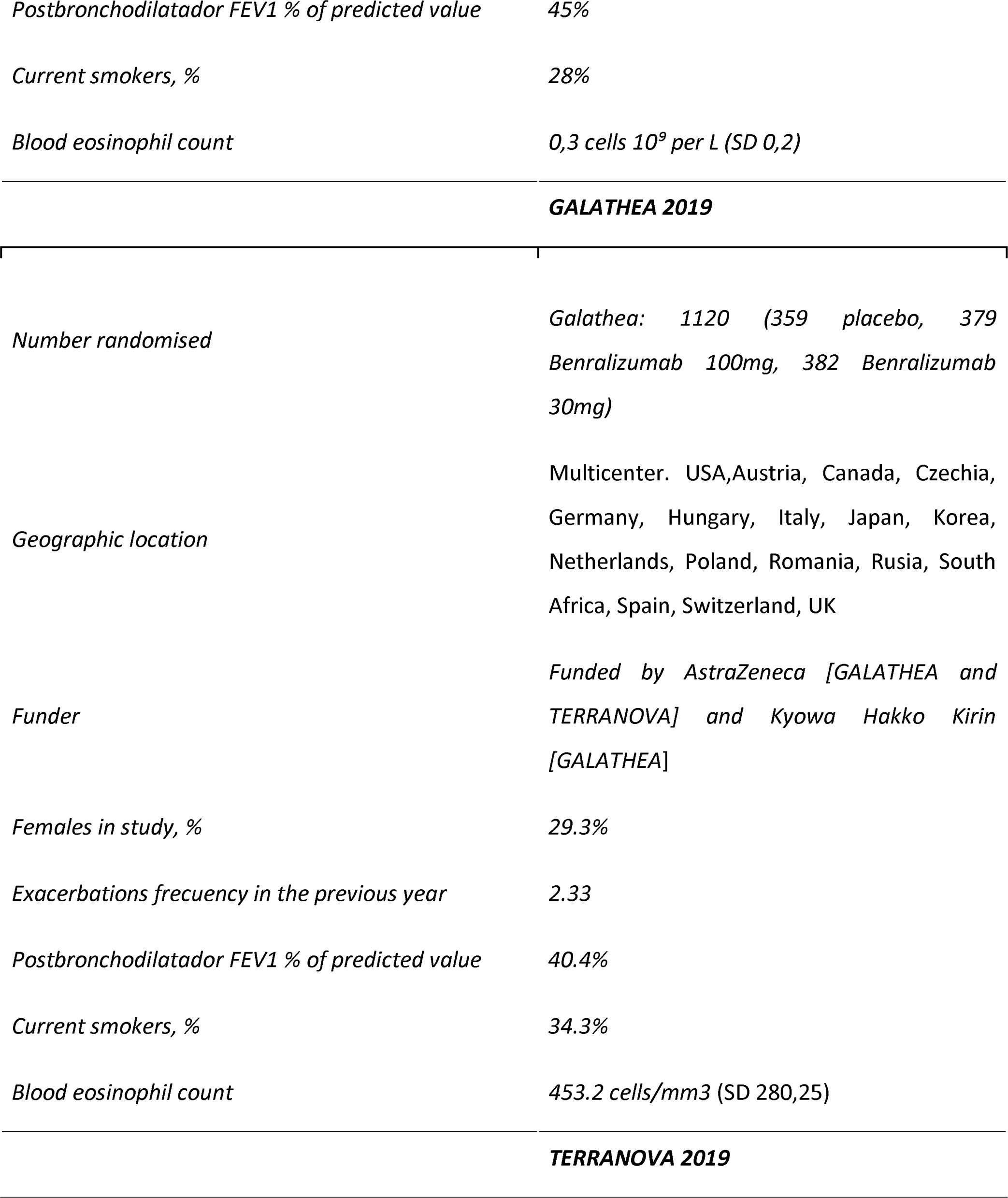
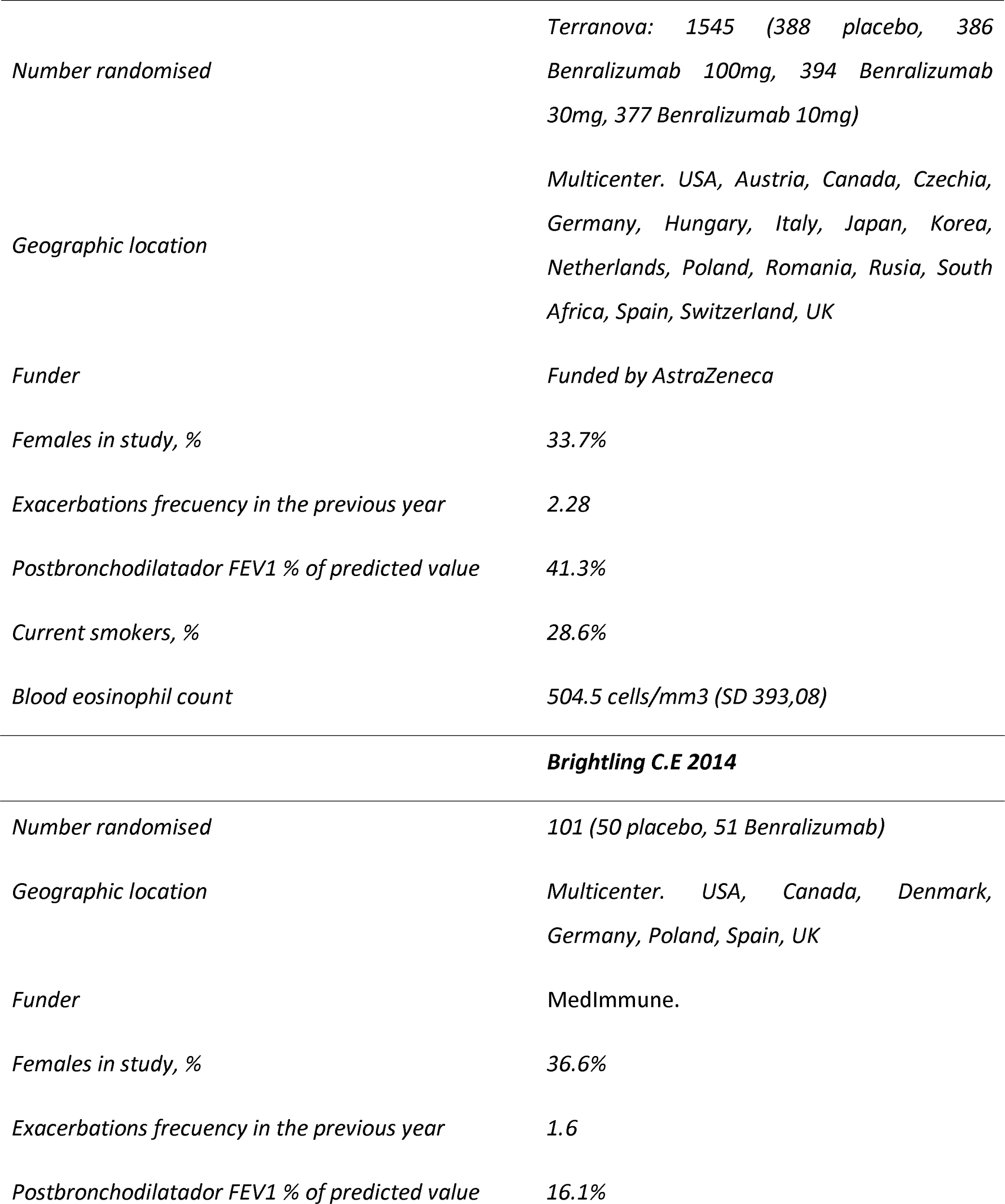
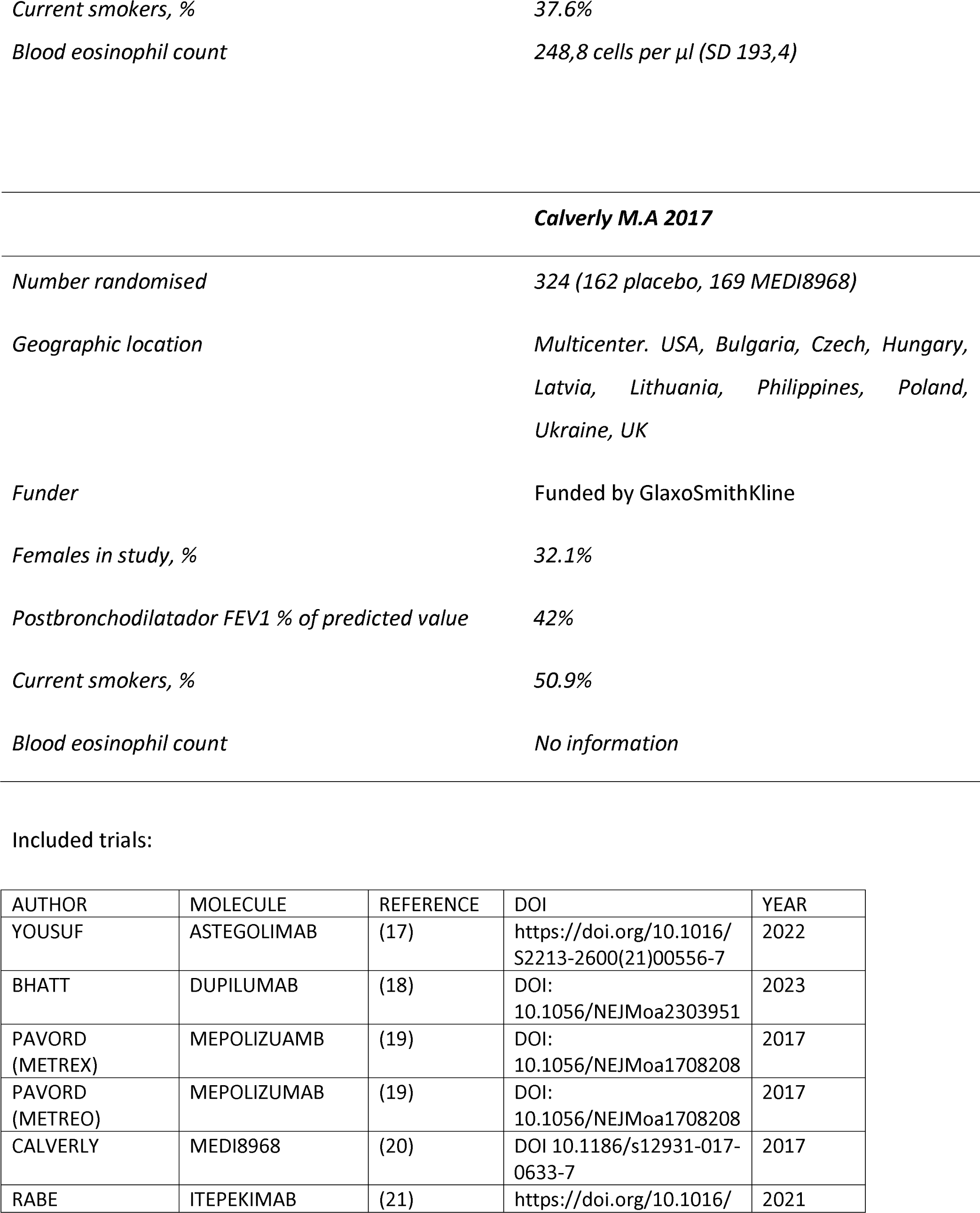
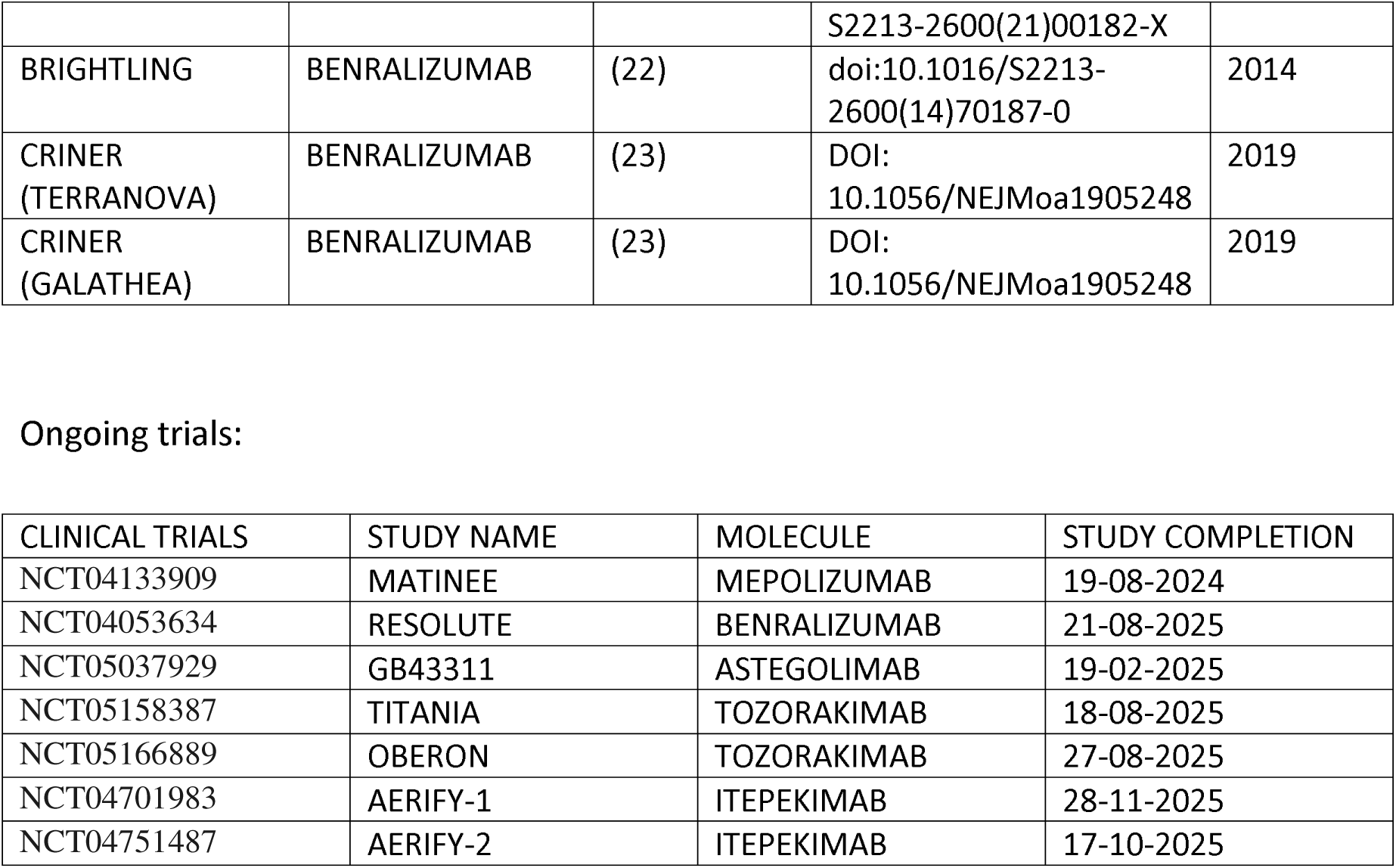

## References

1. Global initiative for chronic obstructive lung disease global strategy for the diagnosis, management, and prevention of chronic obstructive pulmonary disease (2023 report) [Internet]. 2022. Available from: www.goldcopd.org

2. Safiri S, Carson-Chahhoud K, Noori M, Nejadghaderi SA, Sullman MJM, Ahmadian Heris J, et al. Burden of chronic obstructive pulmonary disease and its attributable risk factors in 204 countries and territories, 1990-2019: Results from the Global Burden of Disease Study 2019. The BMJ. 2022;

3. Chen S, Kuhn M, Prettner K, Yu F, Yang T, Bärnighausen T, et al. The global economic burden of chronic obstructive pulmonary disease for 204 countries and territories in 2020–50: a health-augmented macroeconomic modelling study. Lancet Glob Health. 2023 Aug 1;11(8):e1183–93.

4. Connors, A. F., Jr, Dawson, N. V., Thomas, C., Harrell, F. E., Jr, Desbiens, N., Fulkerson, W. J., Kussin, P., Bellamy, P., Goldman, L., & Knaus, W. A. (1996). Outcomes following acute exacerbation of severe chronic obstructive lung disease. The SUPPORT investigators (Study to Understand Prognoses and Preferences for Outcomes and Risks of Treatments). American journal of respiratory and critical care medicine, 154(4 Pt 1), 959–967. 10.1164/ajrccm.154.4.8887592

5. Soler-Cataluña JJ, Martínez-García MÁ, Román Sánchez P, Salcedo E, Navarro M, Ochando R. Severe acute exacerbations and mortality in patients with chronic obstructive pulmonary disease. Thorax. 2005 Nov;60(11):925–31.

6. van Hirtum P V., Sprooten RTM, van Noord JA, van Vliet M, de Kruif MD. Long term survival after admission for COPD exacerbation: A comparison with the general population. Respir Med. 2018 Apr 1;137:77–82.

7. Tashkin DP, Wechsler ME. Role of eosinophils in airway inflammation of chronic obstructive pulmonary disease. Vol. 13, International Journal of COPD. Dove Medical Press Ltd.; 2018. p. 335–49.

8. David B, Bafadhel M, Koenderman L, De Soyza A. Eosinophilic inflammation in COPD: From an inflammatory marker to a treatable trait. Vol. 76, Thorax. BMJ Publishing Group; 2021. p. 188–95.

9. CORDIS EU. research Results. Living Evidence to inform health decisions: Development of capacity for the production and use of reliable, friendly and updated health evidence synthesis. 2023.

10. Moher D, Shamseer L, Clarke M, Ghersi D, Liberati A, Petticrew M, et al. Preferred reporting items for systematic review and meta-analysis protocols (PRISMA-P) 2015 statement. Revista Espanola de Nutricion Humana y Dietetica. 2016;20(2):148–60.

11. Bendersky J, Auladell-Rispau A, Urrútia G, Rojas-Reyes MX. Methods for developing and reporting living evidence synthesis. J Clin Epidemiol [Internet]. 2022 Dec 1;152:89–100. Available from: 10.1016/j.jclinepi.2022.09.020

13. Guyatt GH, Oxman AD, Vist GE, Kunz R, Falck-Ytter Y, Alonso-Coello P, et al. GRADE: an emerging consensus on rating quality of evidence and strength of recommendations. BMJ [Internet]. 2008 Apr 24 [cited 2024 May 19];336(7650):924–6. Available from: https://www.bmj.com/content/336/7650/924

14. Guyatt GH, Oxman AD, Santesso N, Helfand M, Vist G, Kunz R, et al. GRADE guidelines: 12. Preparing Summary of Findings tables - Binary outcomes. J Clin Epidemiol [Internet]. 2013 Feb 1 [cited 2024 May 19];66(2):158–72. Available from: http://www.jclinepi.com/article/S0895435612000327/fulltext

15. Guyatt GH, Thorlund K, Oxman AD, Walter SD, Patrick D, Furukawa TA, et al. GRADE guidelines: 13. Preparing Summary of Findings tables and evidence profiles - Continuous outcomes. J Clin Epidemiol [Internet]. 2013 Feb 1 [cited 2024 May 19];66(2):173–83. Available from: http://www.jclinepi.com/article/S0895435612002405/fulltext

16. Donovan T, Milan SJ, Wang R, Banchoff E, Bradley P, Crossingham I. Anti-IL-5 therapies for chronic obstructive pulmonary disease. Vol. 2020, Cochrane Database of Systematic Reviews. John Wiley and Sons Ltd; 2020.

17. Ohnishi H, Eitoku M, Yokoyama A. A systematic review and integrated analysis of biologics that target Type 2 inflammation to treat COPD with increased peripheral blood eosinophils. Heliyon. 2022 Jun 1;8(6).

18. Yousuf AJ, Mohammed S, Carr L, Yavari Ramsheh M, Micieli C, Mistry V, et al. Astegolimab, an anti-ST2, in chronic obstructive pulmonary disease (COPD-ST2OP): a phase 2a, placebo-controlled trial. Lancet Respir Med. 2022 May 1;10(5):469–77.

19. Bhatt SP, Rabe KF, Hanania NA, Vogelmeier CF, Cole J, Bafadhel M, et al. Dupilumab for COPD with Type 2 Inflammation Indicated by Eosinophil Counts. New England Journal of Medicine. 2023 Jul 20;389(3):205–14.

20. Pavord ID, Chanez P, Criner GJ. Mepolizumab for eosinophilic chronic obstructive pulmonary disease. Journal of Clinical Outcomes Management. 2018 Jan 1;25(1).

21. Calverley PMA, Sethi S, Dawson M, Ward CK, Finch DK, Penney M, et al. A randomised, placebo-controlled trial of anti-interleukin-1 receptor 1 monoclonal antibody MEDI8968 in chronic obstructive pulmonary disease. Respir Res. 2017 Aug 9;18(1).

22. Rabe KF, Celli BR, Wechsler ME, Abdulai RM, Luo X, Boomsma MM, et al. Safety and efficacy of itepekimab in patients with moderate-to-severe COPD: a genetic association study and randomised, double-blind, phase 2a trial. Lancet Respir Med. 2021 Nov 1;9(11):1288–98.

23. Brightling CE, Bleecker ER, Panettieri RA, Bafadhel M, She D, Ward CK, et al. Benralizumab for chronic obstructive pulmonary disease and sputum eosinophilia: A randomised, double-blind, placebo-controlled, phase 2a study. Lancet Respir Med. 2014 Nov 1;2(11):891–901.

24. Criner GJ, Celli BR, Brightling CE, Agusti A, Papi A, Singh D, et al. Benralizumab for the Prevention of COPD Exacerbations. New England Journal of Medicine. 2019 Sep 12;381(11):1023–34.

